# Spatialized Epidemiological Forecasting applied to Covid-19 Pandemic at Departmental Scale in France

**DOI:** 10.1101/2021.11.03.21265855

**Authors:** Matthieu Oliver, Didier Georges, Clémentine Prieur

**Author notes:** Institute of Engineering and Management, Univ. Grenoble Alpes. Email addresses:* (Matthieu Oliver), (Didier Georges), (Clémentine Prieur).

## Abstract

In this paper, we present a spatialized extension of a SIR model that accounts for undetected infections and recoveries as well as the load on hospital services. The spatialized compartmental model we introduce is governed by a set of partial differential equations (PDEs) defined on a spatial domain with complex boundary. We propose to solve the set of PDEs defining our model by using a meshless numerical method based on a finite difference scheme in which the spatial operators are approximated by using radial basis functions. Such an approach is reputed as flexible for solving problems on complex domains. Then we calibrate our model on the French department of Isère during the first period of lockdown, using daily reports of hospital occupancy in France. Our methodology allows to simulate the spread of Covid-19 pandemic at a departmental level, and for each compartment. However, the simulation cost prevents from online short-term forecast. Therefore, we propose to rely on reduced order modeling tools to compute short-term forecasts of infection number. The strategy consists in learning a time-dependent reduced order model with few compartments from a collection of evaluations of our spatialized detailed model, varying initial conditions and parameter values. A set of reduced bases is learnt in an offline phase while the projection on each reduced basis and the selection of the best projection is performed online, allowing short-term forecast of the global number of infected individuals in the department.

## 1. Introduction

Over the past year, the spread of Covid-19 has had more serious consequences than expected. This pandemic has brought about massive organizational changes in many countries, with repeated lockdowns and the use of remote solutions for work, study and personal life. It has also had massive impacts on hospital care services with the need to expand hospital capacity and postpone operations for non-Covid-19 patients. Finally, the pandemic has killed several million people around the world (see, e.g., Pifarré i Arolas et al., 2021). One of the most difficult specifics to model regarding the spread of Covid-19 is the amount of asymptomatic infected individuals, as described, for example, in Al-Tawfiq (2020), which makes the pandemic difficult to track and has led to massive testing campaigns to assess the evolution of the pandemic and the effectiveness of health measures. The efficiency of different strategies of lockdown and testing has been studied (see, e.g., Berger et al., 2020). The incubation period of more than 10 days resulted in a lag between the implementation of restriction rules and their effect on hospital admissions, and made the effectiveness of restrictions more difficult to measure. Forecasting the number of infections and modeling hospital needs is critical to pandemic management and has been the subject of numerous articles over the past year.

Since the start of the epidemic, many studies have been carried out for modeling and forecasting the spread Covid-19. The asymptomatic infected individuals were typically taken into account by building compartmental models that are more detailed than the usual susceptible-infected-recovered (SIR) model (see, e.g., Liu et al., 2021; Berger et al., 2020; Roques et al., 2020; Charpentier et al., 2020, and references therein). These works focus specifically on the effect of lockdown on the propagation of Covid-19 by taking into account detected and undectected infected individuals as well as hospitalizations. Forecasting the number of infections of Covid-19 has been studied in Liu et al. (2021) by using a 3-step approach on the transmission rate during a pandemic outbreak with linear, then exponential increase in infections and finally an exponential decrease of the transmission rate.

In this paper, we propose a spatialized version of a detailed compartmental model inspired by Charpentier et al. (2020), we then restrict the spatial domain to the French department of Isère and propose a calibration procedure over the period of the first French confinement, extending the whole methodology exposed in Guan et al. (2020). This procedure can be adapted to any scale, provided that corresponding data is available. It allows to reproduce the macroscopical behaviour of Covid-19 pandemic on different geographical areas. A preliminary step to the calibration and a second contribution of this paper is the simulation of the model. Our model is defined as a set of spatio-temporal partial differential equations over a complex domain, namely the French department of Isère in our study. We propose a meshless method based on a finite difference scheme relying on radial basis function approximation of the spatial operators (the RBF-FD method, see Fornberg and Flyer (2015)) to solve it, as such an approach is known to be flexible to solve problems on complex domains. To the best of our knowledge, it is the first time RBF-FD is applied in the framework of epidemiology to study the spatio-temporal spread of a disease. Our methodology allows to simulate the spread of Covid-19 pandemic at a departmental level, and for each compartment.

However the simulation cost prevents from online short-term forecast. Therefore we propose as last contribution of the paper to rely on reduced order modeling tools introduced in Bakhta et al. (2021) to compute short-term forecasts of infection number. The strategy consists in learning a time-dependent reduced order model with few compartments from a collection of evaluations of our spatialized detailed model, by varying initial conditions, initial times and parameter values. A set of reduced bases is learnt in an offline phase while the projection on each reduced basis and the selection of the best projection is performed online, allowing short-term forecast of the global number of infections in the department.

The paper is organized as follows. In Section 2, we present an extension of the classical SIR epidemiological model that models the infections, recoveries and hospitalizations spatially over a given geographical area of interest. In Section 3, we present the meshless RBF-FD scheme for solving the set of differential equations from model proposed in Section 2. In Section 4 the model is calibrated using hospital data from Covid-19 outbreak in the department of Isère, France. In Section 5, we use model order reduction techniques to build a surrogate model and extrapolate the number of infected individuals on a 10-day horizon.

## 2. Spatialized *SIRHUD*^+/−^ model

In this section, we present the usual susceptible-infected-recovered (SIR) compartmental model, then we add compartments to adapt to Covid-19 specificities. Finally, we propose an extension of this detailed model to account for the spatial spread of Covid-19 pandemic on a geographical setting, in our framework the Isère department in France.

### 2.1. Usual SIR model

The model presented in this paper is an extension of the usual SIR epidemiological model from Brauer and Castillo-Chavez (2001). The typical SIR model describes a population of constant size, where each individual can be either Susceptible *S* to a disease, Infected *I* with the disease or Recovered *R* from the disease. The transmission of the disease is ruled by the following equations:

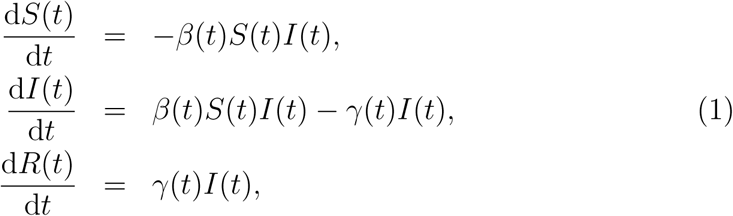

with

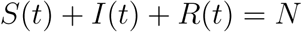

where *β* is the transmission rate of the disease and *γ* the recovery rate.

### 2.2. The SIRHUD^+/−^ model

More detailed models such as the *SIRHUD*^+/−^ model in Charpentier et al. (2020) can be better suited for modeling Covid-19 pandemic as they account for detected and undetected infected individuals *I*^+^ and *I*^−^, detected and undetected recovered individuals *R*^+^ and *R*^−^ as well as hospitalized individuals *H*, individuals receiving intensive care *U* and deceased individuals *D*. This approach is particularly useful in the case of Covid-19 because undetected infections and hospital saturation both play a key role in the handling of the pandemic.

The flow diagram of this model is sketched out in Figure 1.

**Figure 1:**
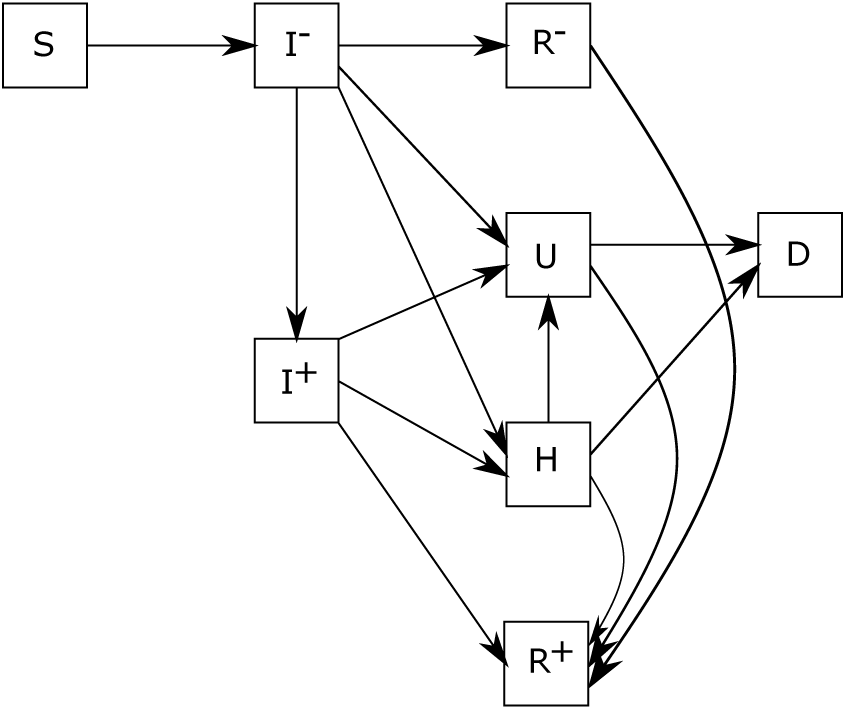
Flow diagram of the pandemic model described by the set of equations in (2)

The dynamics of each compartment is modelled by the following equations:

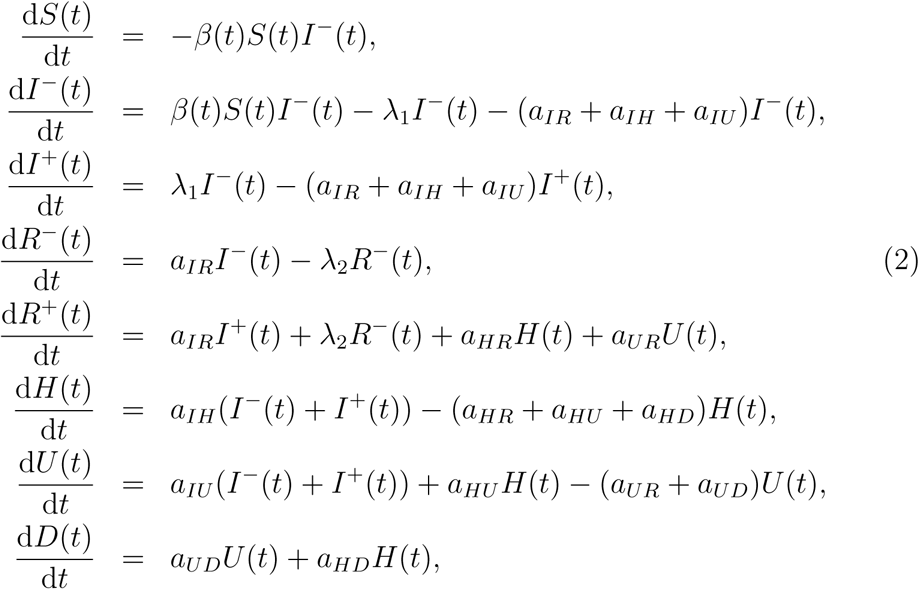

with

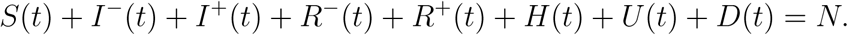

This model was calibrated in Guan et al. (2020) to fit the Covid-19 outbreak at the regional scale in France. In this paper, we aim to enrich this model to better account for spatial non-homogeneity of Covid-19 propagation.

### 2.3. Spatialization of the SIRHUD^+/−^ model

In our model, we keep the same variables as in the *SIRHUD*^+/−^ model from Charpentier et al. (2020), but each variable will be depending on space and time. The governing equations of our model are then:

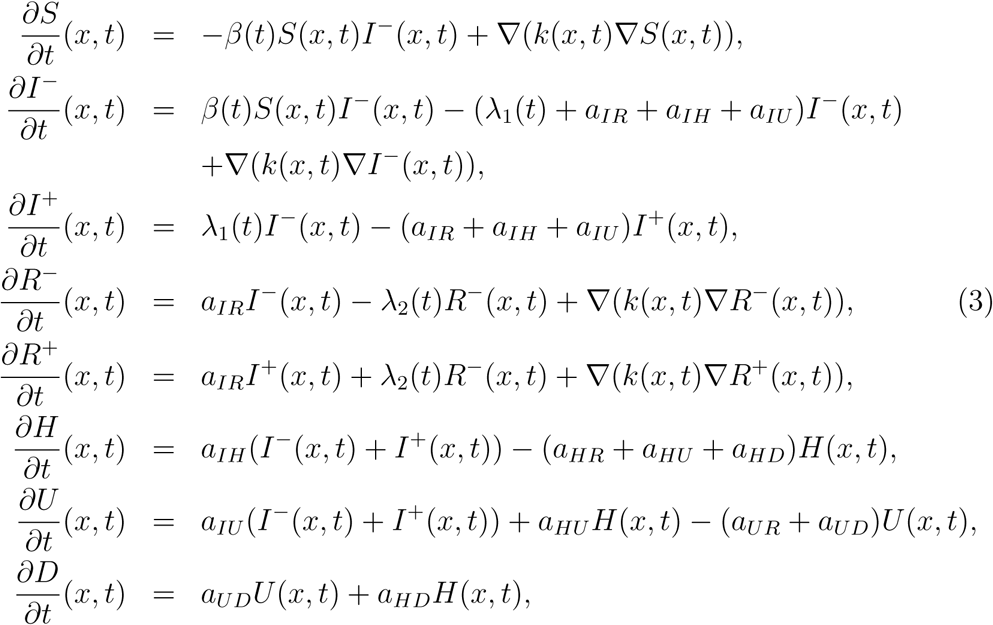

where ∇ denotes the spatial gradient operator.

Note that the variables *S, I*^−^, *R*^−^ and *R*^+^ now feature a diffusion term. This diffusion term translates the fact that individuals from these compartments move locally around their position, whereas individuals from compartments *I*^+^, *H, U* and *D* are isolated and do not transmit the disease. The amount of local movement is quantified by the variable *k*(*x, t*) that may depend on the position *x* and vary in time due to changes in sanitary measures. The model that we choose for the diffusion term is the following:

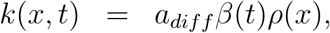

where *ρ*(*x*) is the local population density, *β*(*t*) the transmission rate and *a*_*diff*_ a positive constant. It seems indeed reasonable to assume that in high population density, more local movement occurs, and when sanitary measures are taken, individuals reduce their local movements. The variable *a*_*diff*_ will be part of the calibration variables.

Parameters *a*_*AB*_ are derived from the probability of moving from compartment *A* to *B* and the average duration in compartment *A*. These variables are more precisely described in Guan et al. (2020). Parameter *λ*_1_(*t*) corresponds to the proportion of undetected infected *I*^−^ that are being tested positive, hence moving to compartment *I*^+^, this models virological testing such as PCR tests. *λ*_2_(*t*) corresponds to serological testing of recovered people from *R*^−^ that are moved to *R*+ when tested. These parameters are time-dependent because the testing policies and capabilities have changed during the pandemic. Moreover they can also be used for optimal control, because individuals from *I*^+^ isolate themselves and do not spread the virus. In this work, we considered that the serological testing was negligible: *λ*_2_(*t*) = 0. For the calibration presented in Section 4, virological testing was set to *λ*_1_(*t*) = 0.01 because there was very few testing during the period on which we calibrate the model. However, in the forecasting part of the study that is described in Section 5, this parameter is varied over a wide range of values to replicate different phases of the testing policies and capabilities. In this model, we do not account for the loss of immunity (which could be done by adding a transition from the recovered compartment to the susceptible one), this is because our modeling is done at scale of days to a couple of months, at which scale the loss of immunity is neglected.

## 3. A meshless approach for solving partial differential equations

In this work, we used the radial basis function finite difference (RBF-FD) method from Fornberg and Flyer (2015) to solve the set of partial differential equations defined by (3) in Section 2. To the best of our knowledge, this is the first implementation of the RBF-FD approach to simulate an epidemiological model. Meshless approaches are particularly well-suited to our framework because the domain on which we solve PDE’s is a geographical area that has a complex boundary. In the following, we take *N*_d_ points denoted 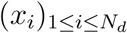 in the interior of the domain. We also take *N*_b_ points denoted 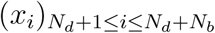 over the boundary. In the following of this section, we focus on compartment of susceptible individuals *S*. Note that all the compartments involved in (3) are treated in a similar way.

### 3.1. Radial Basis Functions

Radial basis functions (RBF) are functions *ϕ*(*x*_*k*_, *x*_*l*_) that depend on the distance *r* = ‖*x*_*k*_ − *x*_*l*_‖ between *x*_*k*_ and *x*_*l*_. We will use the set of points 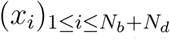 to get an approximation of variable *S*. For each point *x*_*i*_ we denote *I*_*n*_(*x*_*i*_) the indices of the *n* nearest neighbors of *x*_*i*_ in 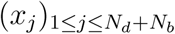. The set of points indexed by *I*_*n*_(*x*_*i*_) defines the RBF-FD stencil of *x*_*i*_. A local approximation of *S* around *x*_*i*_ can then be obtained from the radial basis functions *ϕ*_*k*_(*x*) = *ϕ*(*x, x*_*k*_) *for k* ∈ *I*_*n*_(*x*_*i*_). In this paper, to compute the approximation *S*_*a*_ of *S*, we used the inverse multi quadratic functions:

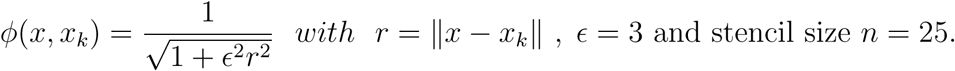

Equation (4) below gives a local approximation of *S* around *x*_*i*_:

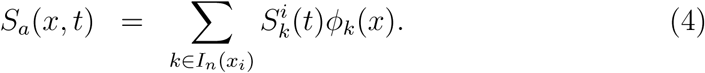

Similarly, we denote *X*_*a*_ the approximation of *X* ∈{*I*^−^, *R*^−^, *R*^+^} whose dynamic is governed by reaction-diffusion PDEs of system (3).

### 3.2. Radial Basis Function Finite Differences

The approximation of the variables around stencil points allows us to locally discretize the diffusion operators that appears in the set of equations (3).

#### 3.2.1. Operator discretization on the domain

Using (4), we can approximate the diffusion operator for variable *S* appearing in (3) as follows:

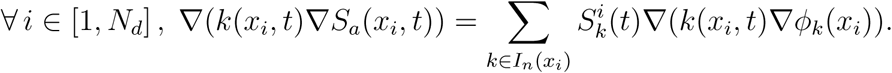

The sum can be written in matrix form

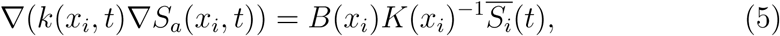

with *B*(*x*_*i*_) containing the operator values at points in the stencil of *x*_*i*_, indexed by *I*_*n*_(*x*_*i*_) = {*j*_1_, …, *j*_*n*_},

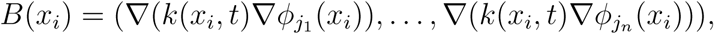

*K* a *n* × *n* symmetric matrix containing the values of the radial basis functions at each couple of points in the stencil of *x*_*i*_,

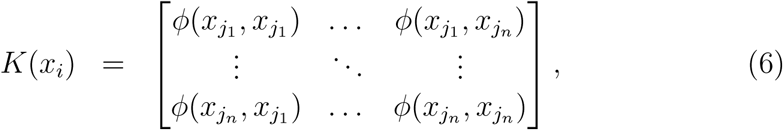

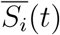 a vector containing the values of *S*_*a*_ at each point of the stencil of *x*_*i*_:

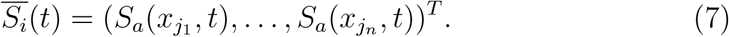

The same diffusion operator approximations as given by (5) hold for the compartment variables *X* ∈{*I*^−^, *R*^−^, *R*^+^} governed by reaction-diffusion PDEs of system (3).

#### 3.2.2. Discretization of the boundary conditions

Boundary conditions can be defined by operator *B*_*c*_, leading for variable *S* to:

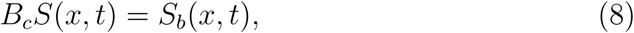

where *S*_*b*_(*x, t*) is the boundary term that is imposed on point *x*. Applying the same method as in the previous section to the boundary points, we obtain for points 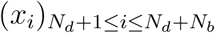 of the boundary:

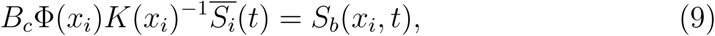

where *K*(*x*_*i*_) and 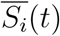 are defined by (6) and (7) respectively, and Φ(*x*_*i*_) is the vector containing the values of the radial basis functions between *x*_*i*_ and the *n* nearest neighbors of *x*_*i*_: Φ(*x*_*i*_) = (*ϕ*_*j*1_ (*x*_*i*_), …, *ϕ*_*j*n_ (*x*_*i*_)). Similar boundary operator approximations as given by (9) hold for other compartment variables *X* ∈ {*I*^−^, *R*^−^, *R*^+^} governed by reaction-diffusion PDEs of system (3).

#### 3.3. Reducing the equations to a system of ordinary differential equations (ODEs)

We finally apply (5) to the diffusion terms in (3) which leads, for points 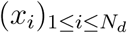 in the interior of the domain, to:

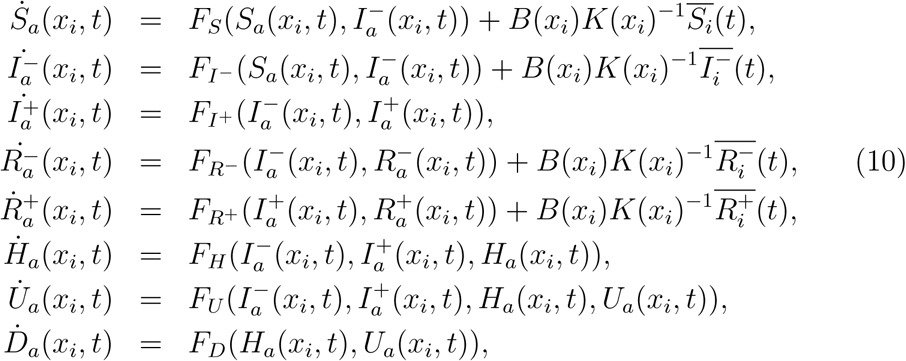

where *F*_*α*_ is the source term of compartment *α* in (3). Additionally, for points *x*_*i*_ on the boundary, the boundary condition for each compartment variable *X* ∈ *{I*^−^, *R*^−^, *R*^+^} can be written as 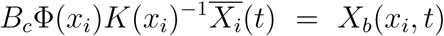. This set of equations with the boundary conditions can be summarized as the following algebraic-differential equations:

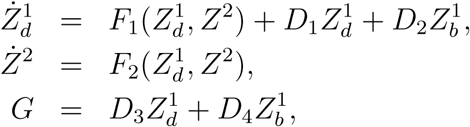

where G is the vector of boundary conditions for the compartment variables at each boundary point:

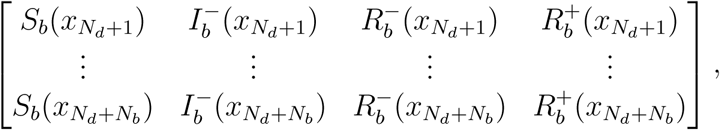

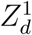 and 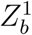 are the vectors of variables *S, I*^−^, *R*^−^, *R*^+^ evaluated at each point of the interior and boundary of the domain respectively, *Z*^2^ is the vector of variables *I*^+^, *H, U, D* evaluated at each of the points (interior and on the boundary) of the domain, and matrices *D*_1_, *D*_2_, *D*_3_, *D*_4_ contain the coefficients of the discretized diffusion and boundary operators using (5) and (9).

When applying Dirichlet conditions as proposed in what follows, we have *D*_3_ = 0 and *D*_4_ = *I*_*d*_, leading to the following system of nonlinear ODEs:

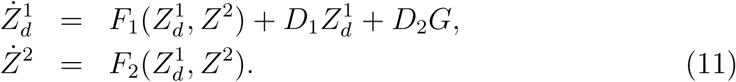

### 3.4. Application to the epidemiological model

The methodology described in the previous section allows us to solve the set of differential equations (3) on a domain given a set of collocation points in the interior of the domain and over its boundary. In our application we want to model the spread of Covid-19 in the department of Isère, France. However our methodology is generic and could be applied to any geographical area of interest, provided corresponding data is available.

#### 3.4.1. Collocation points

For generating the collocation points in the interior of the domain, we used a two-dimensional Halton sequence (see, e.g., Lemieux, 2009) defined on a square and only selected points in the interior of the region, as shown in Figure 2. We thus get a low discrepancy sequence of *N*_*d*_ = 1000 points in the interior of the domain.

**Figure 2:**
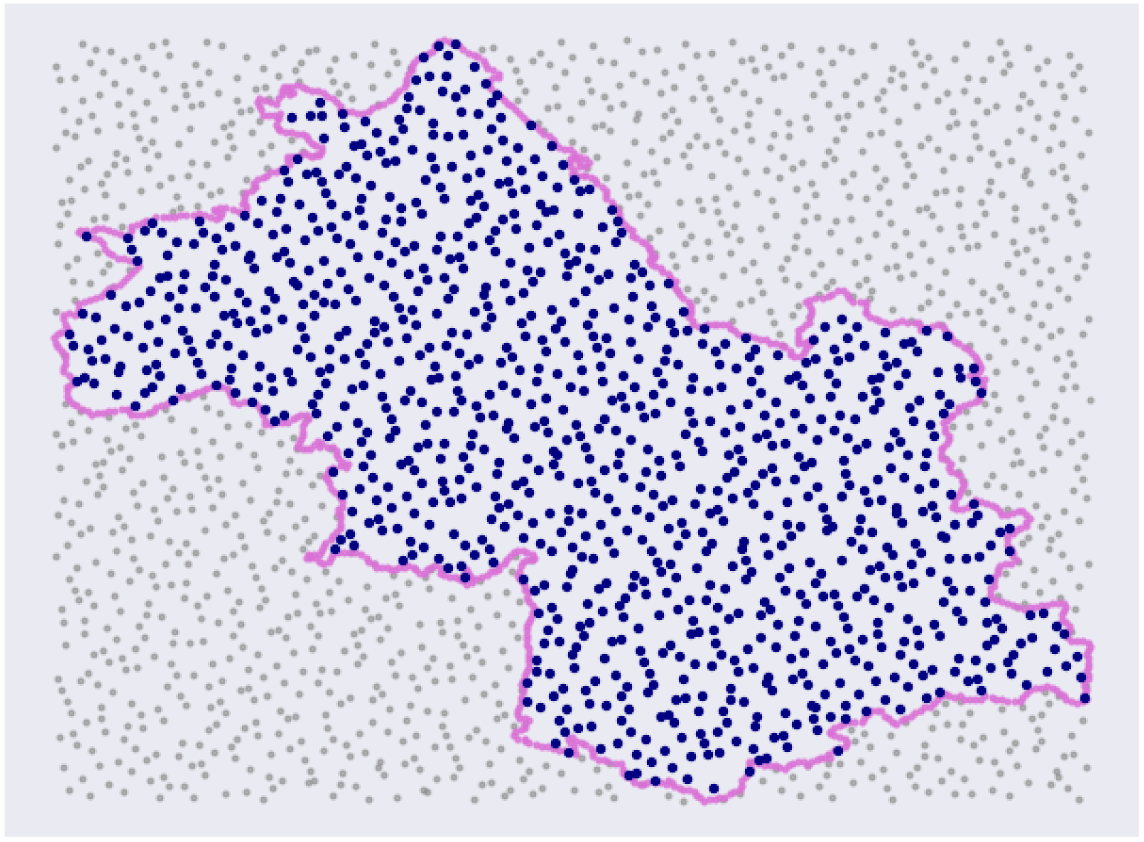
1000 collocation points in the interior of the domain obtained by Halton sequence.

Each collocation point was given the population density value of the closest city taken from INSEE (2014). A polygon describing the boundary of the department was obtained from Lexman (2019) then interpolated to obtain *N*_*b*_ = 800 points along the boundary.

#### 3.4.2. Boundary conditions

As shown in Figure 3, the population density is very heterogeneous over the domain, with the south-east being very rural with densities under 100 *hab/km*^2^ and the north-east and center of the department being a lot more urbanised. We chose to apply null boundary condition on the boundary points *x*_*i*_ with density *ρ*(*x*_*i*_) *< d*_*min*_ = 200 *hab/km*^2^. On the rest of the boundary we assumed that the population of each compartment was equal to the average population of that compartment over the domain. More precisely, for *N*_*d*_ +1 ≤ *i* ≤ *N*_*d*_ +*N*_*b*_ with *ρ*(*x*_i_) ≥ *d*_*min*_, we assumed 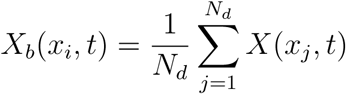.

**Figure 3:**
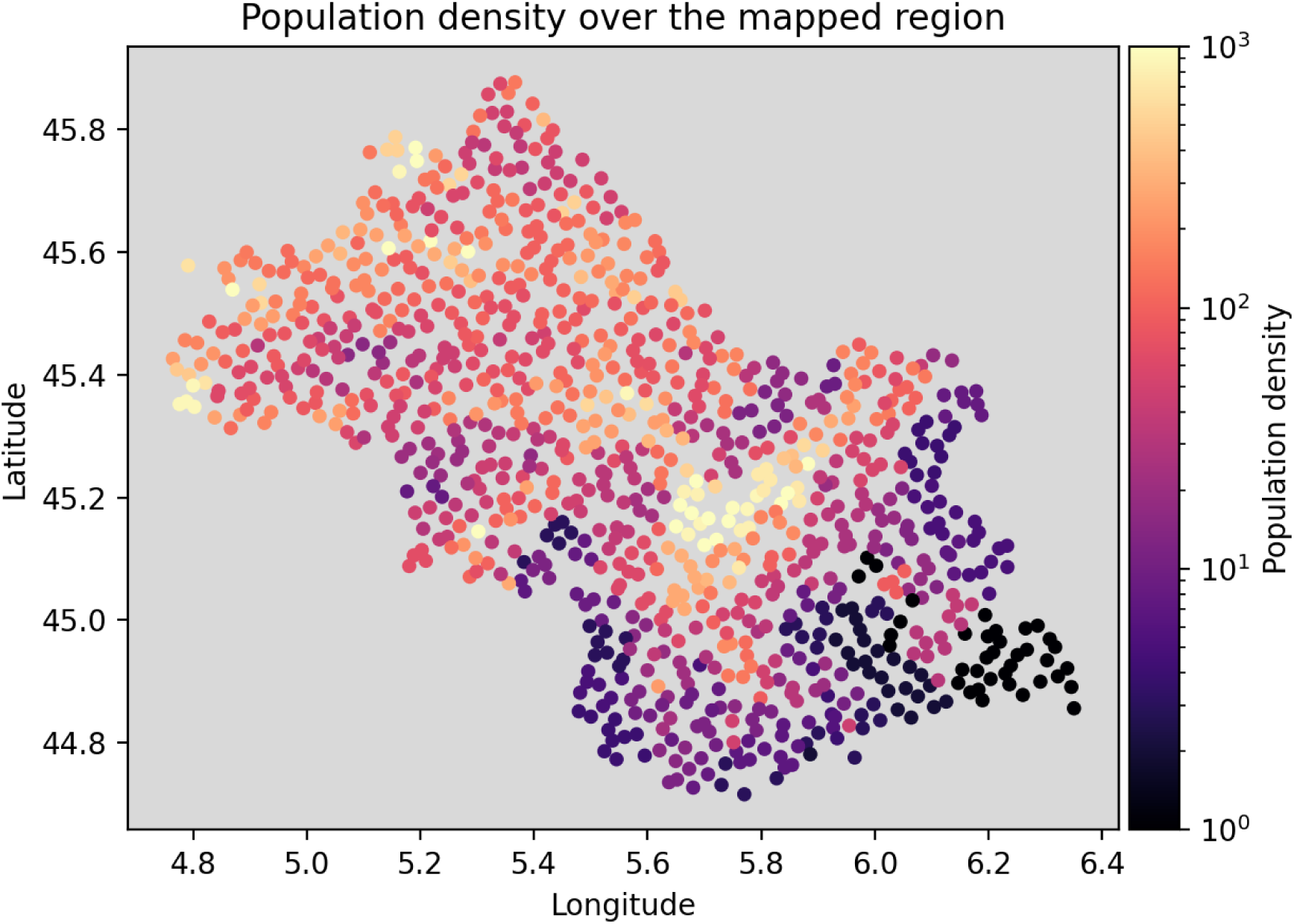
Population density attributed to each collocation point.

#### 3.4.3. Initial Conditions

At *t* = 0, the proportion of individuals in each compartment should be defined in every point of the domain, which represents 8 × *N*_*d*_ = 8000 parameters. These proportions are not known everywhere nor for each compartment and thus have to be calibrated. To reduce the number of parameters to be calibrated, the proportions of infected and hospitalized individuals are modelled by a uniform distribution over the domain. For *i* ≤ *N*_*d*_, we assume:

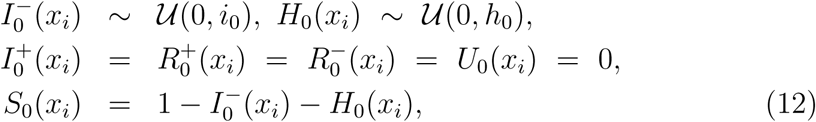

where *i*_0_*/*2 and *h*_0_*/*2 are the initial proportions of infected and hospitalized individuals in the domain.

Then we used a numerical integrator based on backward differentiation formulas to solve the stiff system of ordinary differential equations expressed in (11). The time step was set equal to one day.

## 4. Model Calibration

In this section, we propose a calibration procedure for our spatialized detailed model, extending the procedure proposed in Guan et al. (2020), based on hospital data from Santé Publique France (2021).

### 4.1. Initial calibration

To initialize the calibration procedure of our spatialized model, we took the result of the calibration of its non-spatialized version performed in Guan et al. (2020).

### 4.2. Variables to fit

We used data from Santé Publique France (2021), namely the number of hospitalizations *H*_*obs*_, the number of intensive care hospitalizations *U*_*obs*_, the death toll *D*_*obs*_ and the number of recoveries from hospitalization *R*_*hobs*_. The variables *H, U* and *D* are already involved in the model description (3), we only added to the model the number of returns from hospitalization:

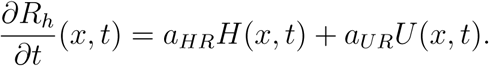

Because of the weekly fluctuations of the data, a rolling average of 7 days was applied to the data from Santé Publique France (2021).

### 4.3. Calibration period

We calibrated our model over the first wave of Covid-19 in France which took place between March 17^th^ and May 11^th^ as was done in Guan et al. (2020). This period is suitable for calibration purposes because the sanitary conditions remained stable over this time period. Moreover, we used hospital data as this data set is the more reliable one, in opposition to test data that is submitted to changes in testing capabilities and policies.

### 4.4. Calibration parameters

Our model has numerous parameters that are presented in this section. Because of the amount of parameters we only calibrated a selection of them, the rest staying at the value given in Guan et al. (2020) as detailed hereafter.

#### 4.4.1. Transmission rate β

The transmission rate is one of the most sensitive parameters to tune the model (see, e.g., Da Veiga et al., 2021), hence its parametrization has to be chosen carefully. We used the one proposed in Liu et al. (2021) and added a non-zero asymptotic value *β*_∞_: *β*(*t*) = (*β*_0_ − *β*_∞_) exp^−*μ*(*t*−*τ*)^ + *β*_∞_. Note that we chose a transmission rate that is only dependent on time. This assumption is motivated by the fact that sanitary measures were the same all around the region of interest. However, for a larger scale implementation of this model, the transmission rate could be defined region-wise depending on the scale at which sanitary measures are enforced.

This model is quite reasonable for lockdown conditions: after time *τ*, the transmission rate decreases exponentially at speed *μ* from it’s original value *β*_0_ to an asymptotic value *β*_∞_. An example can be seen on Figure 4 above. We then have 4 parameters to calibrate *β*.

**Figure 4:**
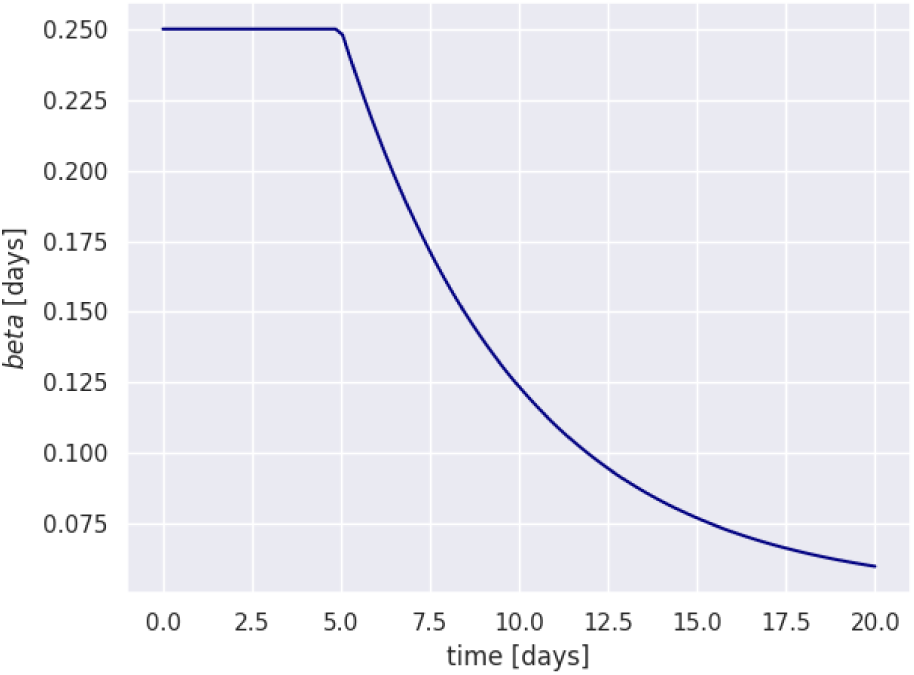
Transmission rate *β* over time with coefficients *β*_0_ = 0.25, *μ* = 0.2, *τ* = 5 and *β*_∞_ = 0.05.

#### 4.4.2. Diffusion coefficient

The diffusion coefficient *a*_*diff*_ appearing in (3) accounts for the non-homogeneity of the transmission of the disease.

#### 4.4.3. Compartment transition variables

The parameters *a*_XY_ appearing in (3) depend on transition times and probabilities. The detailed formulas to obtain *a*_XY_ are written in Guan et al. (2020). The transition times and probabilities coming into play are:

- *p*_*a*_, *p*_*h*_ and *p*_*u*_ the respective probabilities of needing no hospital care, hospitalization and intensive care,
- *p*_*hu*_, *p*_*hd*_, *p*_*ud*_ the respective probabilities of needing intensive care or dying while hospitalized or in intensive care,
- *N*_*a*_ and *N*_*s*_ the recovery times of asymptomatic and lightly symptomatic individuals with no hospitalization,
- *N*_*ih*_, *N*_*hd*_, *N*_*hu*_, *N*_*ud*_, *N*_*hr*_ and *N*_*ur*_ the average transition times between compartments *I*^±^, *H, U* and *D*.

This totals 14 constants to be calibrated to account for recoveries.

#### 4.4.4. Initial Conditions

As detailed in Section 3.4.3, the initial conditions are reduced to the initial proportions *i*_0_*/*2 and *h*_0_*/*2 of *I*^−^ and *H*. If the calibration was done in a later phase of the pandemic, we could add initial proportions of recovered individuals (*R*^±^) as well as intensive care occupation (*U*).

### 4.5. Calibration process

We presented the variables to fit and the corresponding French hospital data, the set of parameter values for the initialization, the time period over which we performed the calibration and finally all the parameters to tune, including initial conditions. In this section we detail the optimization procedure and we present the results of the calibration.

#### 4.5.1. Loss function

As we used hospital data aggregated on the department of Isère to fit our spatialized model, we first need to define *H*_*sim*_, *U*_*sim*_, *D*_*sim*_ and *R*_*hsim*_ the total number of individuals in the department of Isère of the corresponding compartment in the simulation output. We then define the loss function to be minimized over the set of parameters *p* as:

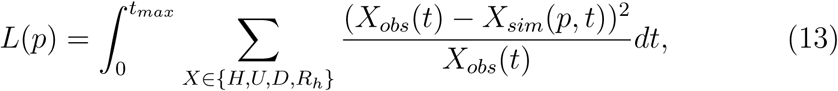

with *t*_*max*_ = 55 *days*, which is length of the first lockdown in France.

#### 4.5.2. Minimization method

For minimizing the calibration loss (13), we used a stochastic optimization Python package noisyopt Mayer (2017). We also gave to the solver bounds for each parameter equal to [*v*_0_*/*5, 5*v*_0_] where *v*_0_ is the nominal value obtained from Section 4.1.

#### 4.5.3. Results

Only a subset of parameters was calibrated. Parameters *p*_*a*_, *p*_*h*_, *p*_*u*_, *p*_*hu*_, *p*_*hd*_, *p*_*ud*_, *N*_*a*_ and *N*_*s*_ were kept to the value obtained in the calibration of the non-spatialized version of our model performed in Guan et al. (2020): *p*_*a*_ = 0.9, *p*_*h*_ = 0.9, *p*_*u*_ = 0.2, *p*_*hu*_ = 0.001, *p*_*hd*_ = 0.176, *p*_*ud*_ = 0.3, *N*_*a*_ = 7.82 and *N*_*s*_ = 15. This last point is commented in Section 4.5.4.

The optimal parameters to fit the data as viewed on Figure 5 are given in Table 1.

**Table 1:** Optimal calibration parameters.

| $\beta_0$ | $\mu$ | $\tau$ | $\beta_\infty$ | $i_0$ | $h_0$ | $N_{ih}$ | $N_{hd}$ | $N_{hu}$ | $N_{ud}$ | $N_{hr}$ | $N_{ur}$ |
| --- | --- | --- | --- | --- | --- | --- | --- | --- | --- | --- | --- |
| 0.257 | 0.109 | 10.3 | 0.00875 | 5.25e-3 | 4e-5 | 9.5 | 1.5 | 0.65 | 1.8 | 1.56 | 1.03 |

**Figure 5:**
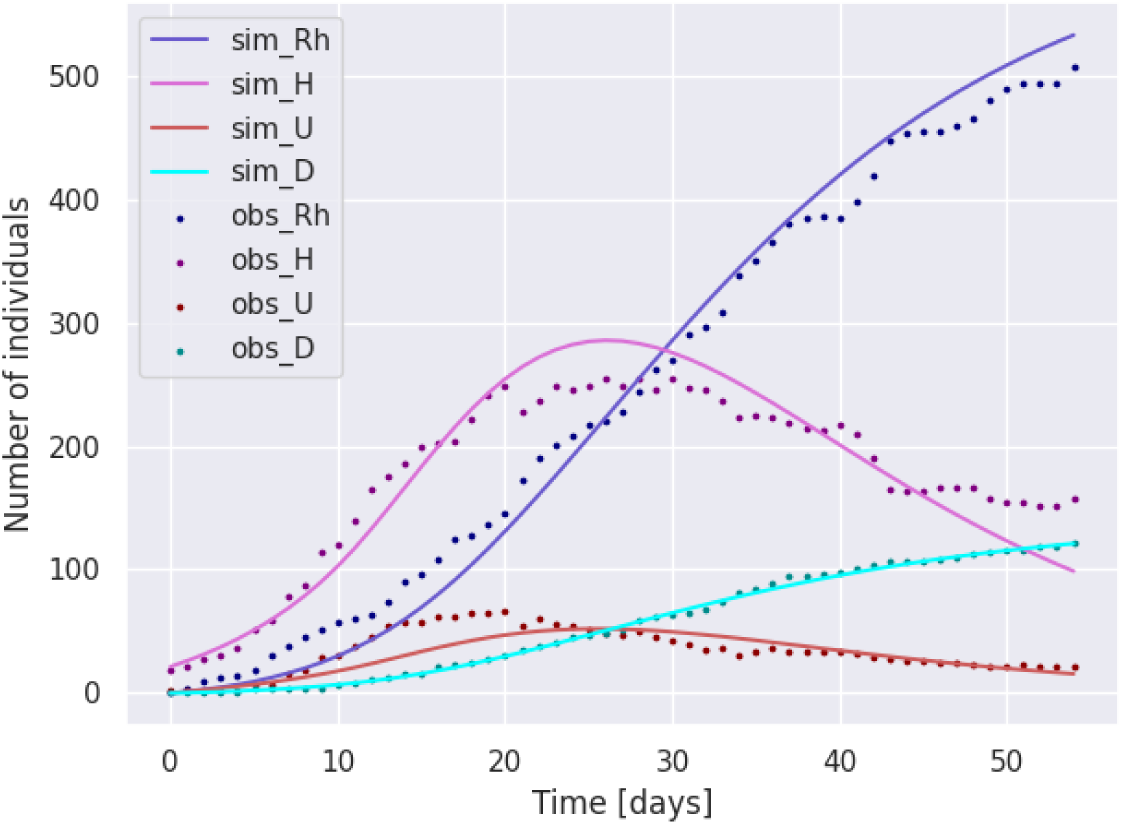
Fitting of compartments *H, U, D* and *R*_*h*_ in Isère during the first lockdown in France (March-May) using the spatialized model described by (3).

#### 4.5.4. Calibration limitations

This calibration was done under lockdown condition where the evolution of *β*(*t*) was shown to be exponentially decreasing Liu et al. (2021). This assumption does not hold in the general case; the evolution of the transmission rate can follow more complex variations as will be shown in the forecasting of *β* that is carried out in Section 5. We also assumed that the probabilities of moving from one compartment to the other and the average duration in each compartment remained constant regardless of the sanitary measures, as being epidemiological parameters. Note however that this assumption may not hold when studying subsequent variants of Covid-19 and their propagation in a partially vaccinated population, hence these parameters should be included in the calibration procedure if studying this later phase of the pandemic.

## 5. Forecasting using reduced order modeling

We saw in Section 4 that our spatialized model is able to reproduce the outbreak of Covid-19 at the scale of the department while giving a very localized information on hospital needs and infection number. However this model is computationally expensive compared to a non-spatialized model, this is why we propose to rely on reduced order modeling to fasten the online computations required for short-term forecasts of the infection number. In this section, we extend the model reduction tools introduced in Bakhta et al. (2021) to the treatment of our spatialized model. In the following of this section, we explain how to build a reduced basis for extrapolating variables 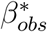 and 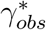 defined in (14). We then detail how it is possible to use the reduced basis to compute online the *n*_*forecast*_-day forecasts of the global infection number in the department of Isère from an observation period of *n*_*calib*_ = 10 days.

### 5.1. Fitting epidemiological data with a time dependent SIR model

In this section, we project our spatialized 8-compartment model into a time-dependent SIR model described by (1). This is done by aggregating the spatial output of each compartment over the domain and regrouping separate compartments in either *S, I* or *R* as follows:

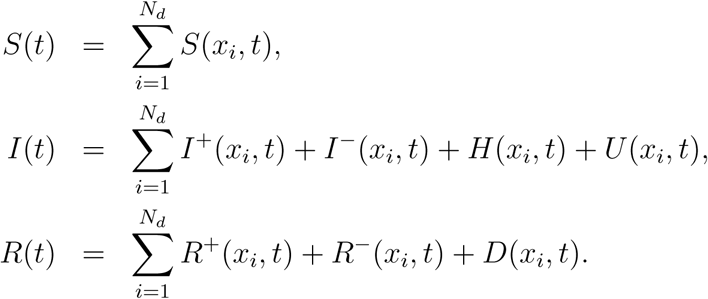

Starting from an initial epidemiological state (*I*(*t* = 0) = *i*_0_, *R*(*t* = 0) = *r*_0_), the evolution of the SIR model is ruled by parameters *β*(*t*) and *γ*(*t*). As a consequence, forecasting the total number of infections and recoveries over the domain can be done by extrapolating *β*(*t*) and *γ*(*t*). Let *S*_*obs*_(*t*), *I*_*obs*_(*t*) and *R*_*obs*_(*t*) denote the evolution of *S*(*t*), *I*(*t*), *R*(*t*) observed over a time period. We then define:

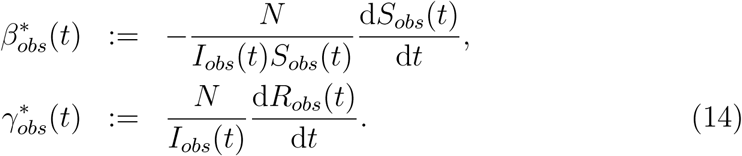

Under mild assumptions that are detailed in Bakhta et al. (2021), equations in (14) allow a simple SIR modeling of the observed data starting from the initial state 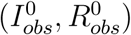. Note that in (14), one can replace observation data by simulation data from a model that outputs *S, I* and *R* over time, leading to the definition of 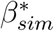 and 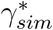 that we will use for forecasting as detailed in Section 5.5.2. It is possible to infer *I*_*obs*_ and *R*_*obs*_ from the data provided by Santé Publique France (2021), applying an adjustment factor (see Bakhta et al. (2021)[Section 4.1] for more details): *I*_*obs*_ = *f H*_*obs*_, *R*_*obs*_ = *f R*_*hobs*_ + *D*_*obs*_, with *f* = 15 computed from the literature (Mizumoto et al., 2020; Di Domenico et al., 2020) and assessing the proportion of individuals needing hospitalization when infected with Covid-19.

The evolution of 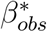 is shown on Figure 6, on which one observes the influence of lockdown periods and sanitary measures. Note that 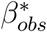 takes negative values around July, 2020. This reflects the very low tendency of hospital data in Isère during the summer of 2020. During that period, the SIR dynamics did not fit well the hospital data, leading to negative values for 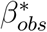. However this only happened in a short interval of time during which the pandemic was stable.

**Figure 6:**
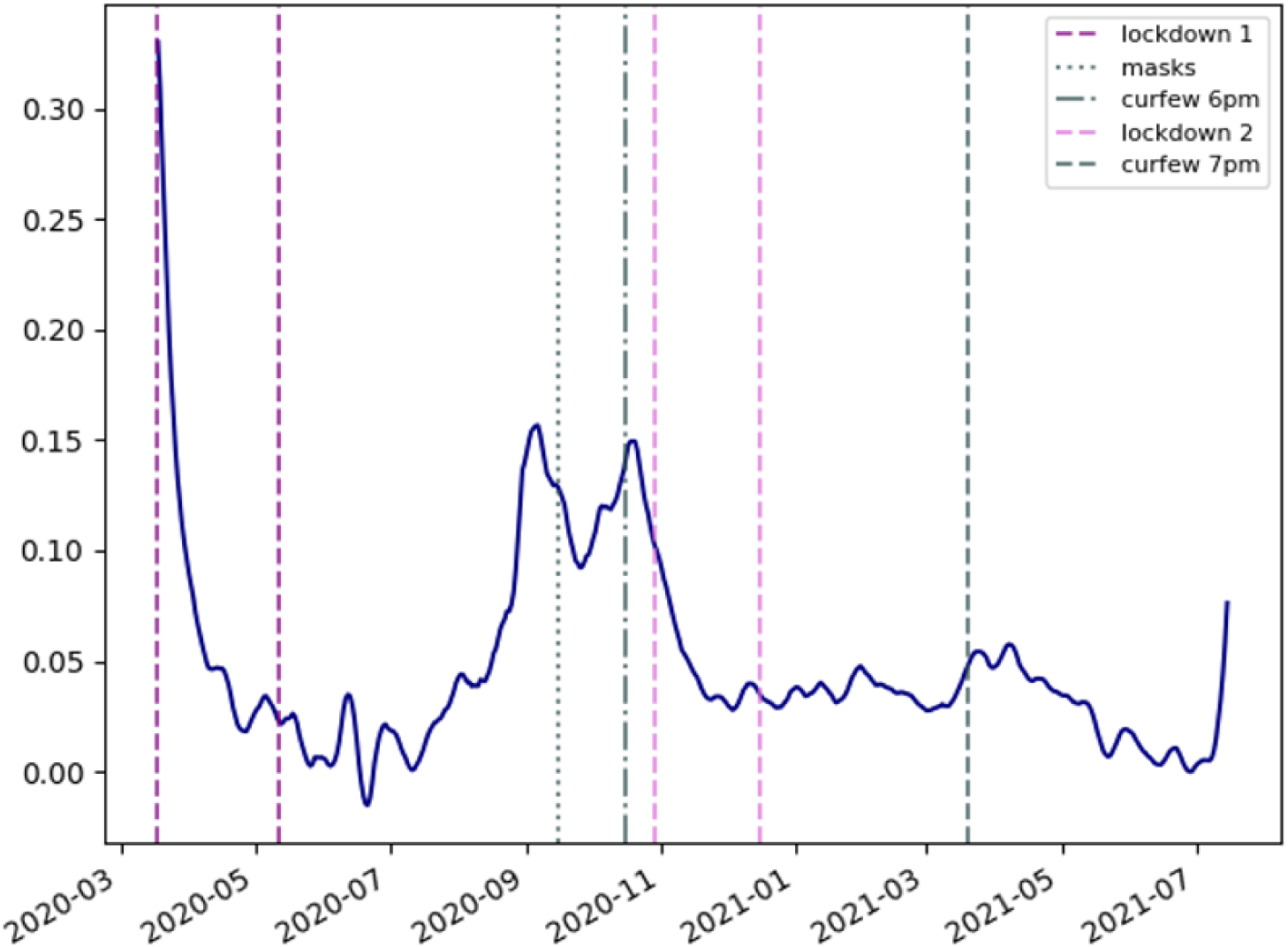
Evolution of 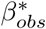 obtained from hospital data between March 2020 and July 2021. We can see that the dates of the sanitary measures match the change of variations of 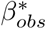 At the very end of the profile we can see the start of the “delta wave” that occured during the redaction of this work.

### 5.2. Detailed model evaluations

In order to fasten the online computations we built a reduced order model. A key stone in the reduction of the model is the construction of a reduced basis from a set of evaluations of the detailed model. We built *K* different reduced bases (ℬ_*k*_)_1≤*k*≤*K*_ for extrapolating the short-term evolution of *β*_*obs*_ and *γ*_*obs*_, by moving the initial time. We then selected the most suited reduced basis and used it for actual forecasting. Note that for the construction of the reduced bases, the detailed model described in (3) was slightly modified by considering that *β* and *λ*_1_ do not depend on time. Then the model was run with different set of initial conditions and parameter values. The range of values we considered for initial conditions and for parameters *β* and *λ*_1_ is provided in Table 2. Even if the bounds for each parameter were chosen to be consistent with the sanitary condition at the start of the second wave of Covid-19 in France (September, 2020), the idea is that the range of admissible values is chosen wide enough to match different calibration periods. Each parameter was sampled from a Halton sequence of length *n*_*runs*_ = 500. The use of a Halton sequence allows to better explore the interval in which each parameter is defined. Other parameters were fixed from Table 1 in Section 4.5.3.

**Table 2:** Bounds for the training set of parameters.

| Parameter | $\beta_0$ | $\lambda_1$ | $i_0^-$ | $i_0^+$ | $r_0^-$ | $r_0^+$ | $h_0$ | $u_0$ | $k_{diff}$ |
| --- | --- | --- | --- | --- | --- | --- | --- | --- | --- |
| Min | 0.05 | 0.1 | 0.03 | 0.0015 | 0.03 | 0.01 | 9e-5 | 8e-6 | 1e-8 |
| Max | 0.15 | 0.5 | 0.06 | 0.0025 | 0.06 | 0.1 | 1.1e-4 | 1.2e-5 | 5e-7 |

Then, the detailed model was run with these *n*_*runs*_ combinations of parameter values over a period of *n*_*eval*_ = 200 days, producing a set of detailed outputs - the number of individuals in each of the 8 compartments over time - that are collapsed into SIR outputs using the following rules:

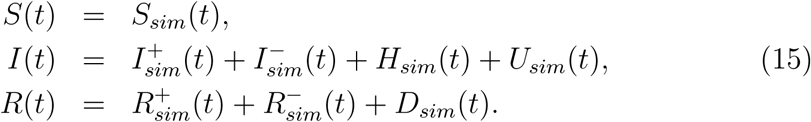

For each run, we computed a realization of 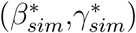 using (14) on the *SIR* outputs. The set of realizations of 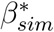 and 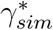 are respectively denoted ***B*** and ***G*** in the following sections. The infection number over time is shown in Figure 7 for a selection of 50 sets of parameter values. In Sections 5.3 and 5.4 we detail the construction of the reduced order model from the set of realizations of 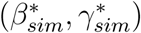.

**Figure 7:**
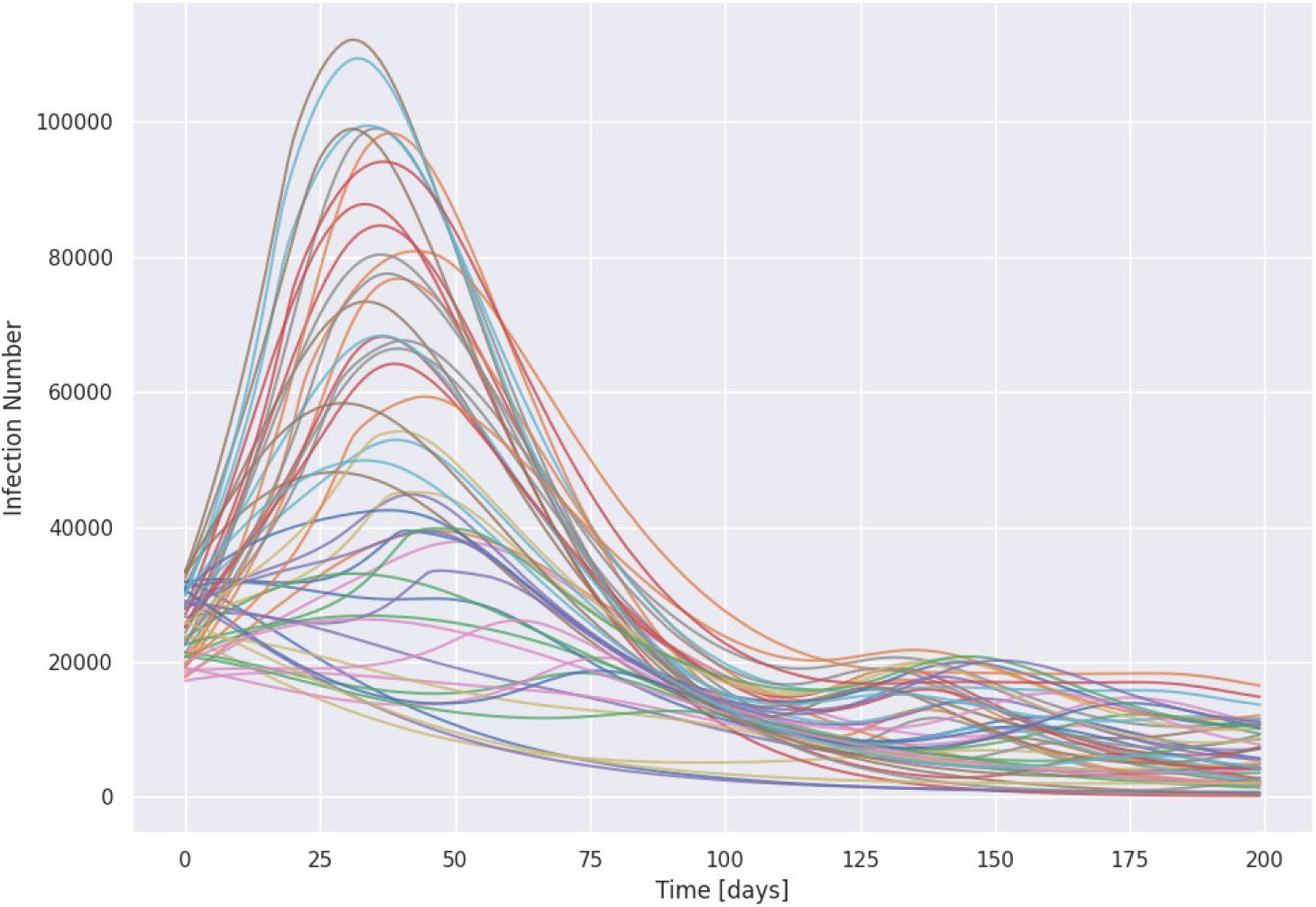
Evolution of the infection number over *n*_*eval*_ days whith 50 sets of parameter values from the training set.

On most of the simulations in Figure 7, we observe a wave growing over 50 days after the start of the simulation, as the bounds in Table 2 were chosen to be coherent with the sanitary conditions in France at the start of the second wave in France (September, 2020). However, recall that we widen the bounds in order to build a more flexible set of reduced bases, which explains why we observe other scenarios, such as simulations with a wave starting around 110 days after the starting time, or even simulations without any wave.

Even though Figure 8 was obtained from detailed model evaluations with *β* not depending on time, the projection to a simple SIR model of the output as described in (15) led to a set of realizations of time-dependent 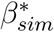.

**Figure 8:**
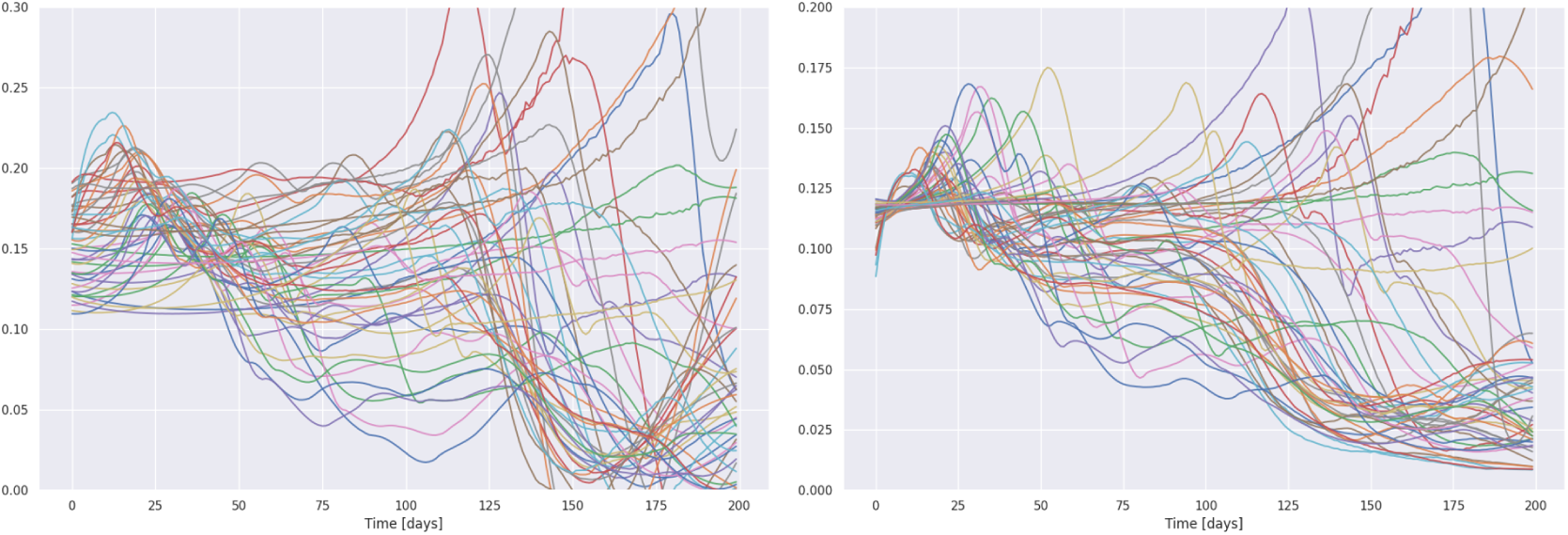
Subsets of ***B*** (left) and ***G*** (right) obtained by applying (14) to the SIR trajectories outputted by the detailed model.

### 5.3. Simulation output reduction

From the set of realizations of 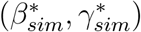 shown in Figure 8 we built respective sets of functions ℬ and 𝒢 defined on *n*_*eval*_ = 200 *days* for *β* and *γ* that can reproduce the typical variations of the detailed model. We used two different methods for building ℬ and 𝒢: one based on Singular Value Decomposition and another one on a greedy algorithm. In the following, we detail the construction of ℬ from ***B***, the same steps were applied to construct 𝒢 from ***G***.

#### 5.3.1. Singular Value Decomposition

This approach consists in discretizing the functions in ***B*** at a 1-day timestep. We then obtain a *n*_*eval*_ × *n*_*runs*_ matrix denoted *M*_*β*_ containing all the 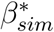 in ***B***. We then compute eigenvalues and eigenvectors of the matrix 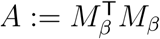. The ordered eigenvalues of *A* are shown in Figure 9. Then ℬ^*SVD*^ is composed with the *n*_*span*_ eigenvectors with the largest eigenvalues. The basis we obtained for *n*_*span*_ = 4 is shown on Figure 10. We remark on Figure 9 that the magnitude of the 10 largest eigenvalues is significantly larger than the rest, which is why with this approach we never used more than 10 functions to build ℬ^*SVD*^ in the following.

**Figure 9:**
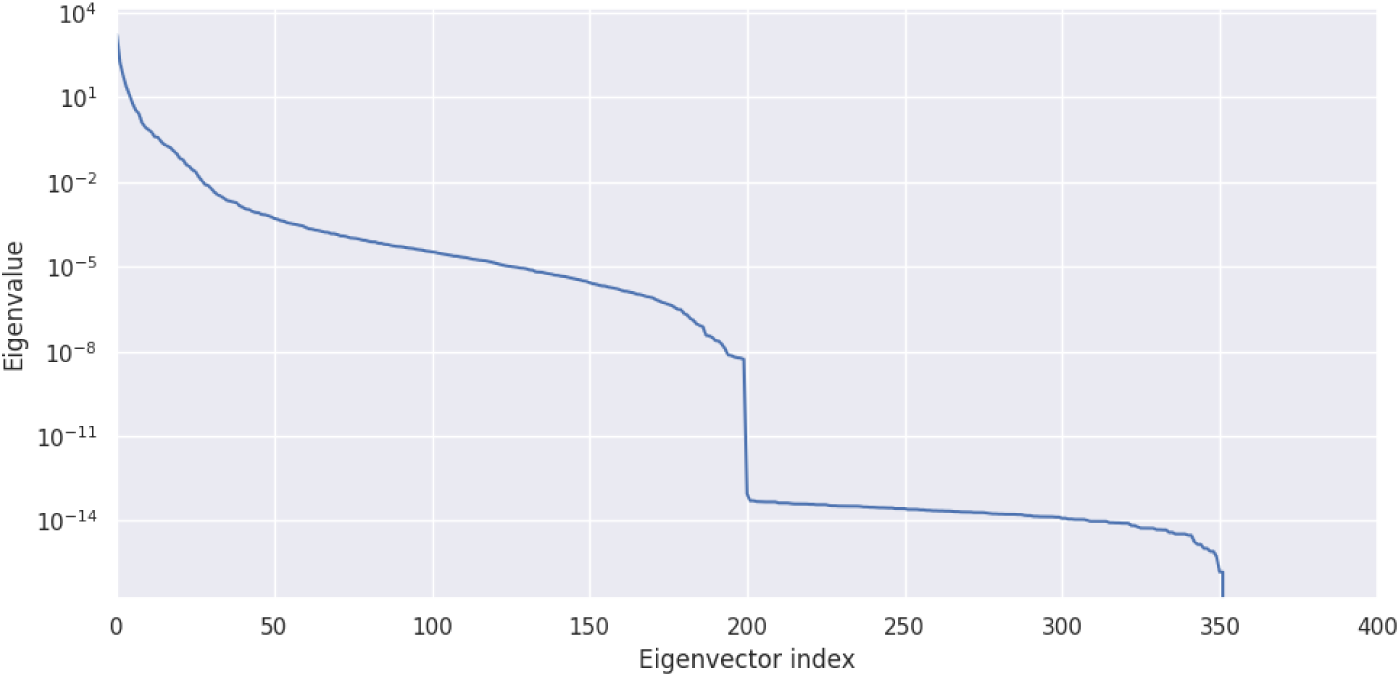
Eigenvalues of matrix *A*.

**Figure 10:**
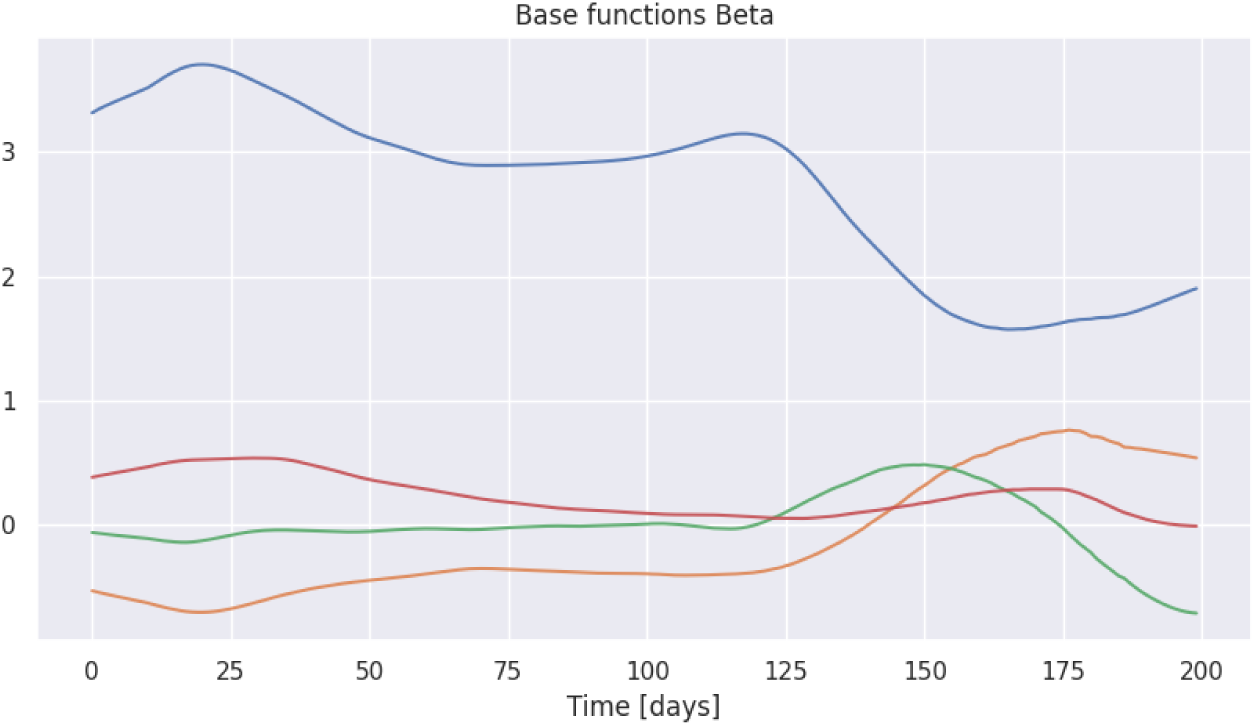
The set ℬ^*SVD*^ of functions obtained through the Spectral Values Decomposition-based algorithm with *n*_*span*_ = 4.

As can be seen on Figure 10, the elements in ℬ^*SVD*^ are not constrained to remain positive. Section 5.3.2 tackles this issue by constructing in a greedy way a basis of positive functions.

#### 5.3.2. Greedy algorithm

The method is based on the algorithm called Enlarged Nonnegative Greedy (ENG) presented in Bakhta et al. (2021). We first use a greedy algorithm to build a set 𝒜 of nonnegative functions selected from ***B***. This set is initialized as follows: 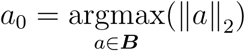.

Then, while 𝒜 has *k* ≤ *n*_*span*_ elements (𝒜 = {*a*_1_, …, *a*_*k*_}), we add the function *a*_*new*_ ∈ ***B*** that has the maximal distance from *Span* (𝒜):

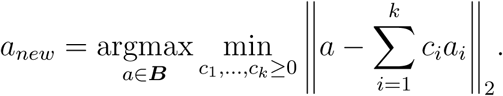

We thus obtain a set of *n*_*span*_ nonnegative elements of ***B*** that will be used to build the set ℬ^*ENG*^ using the enlarge cone algorithm from Bakhta et al. (2021):

##### Algorithm 1: Enlarge Cone

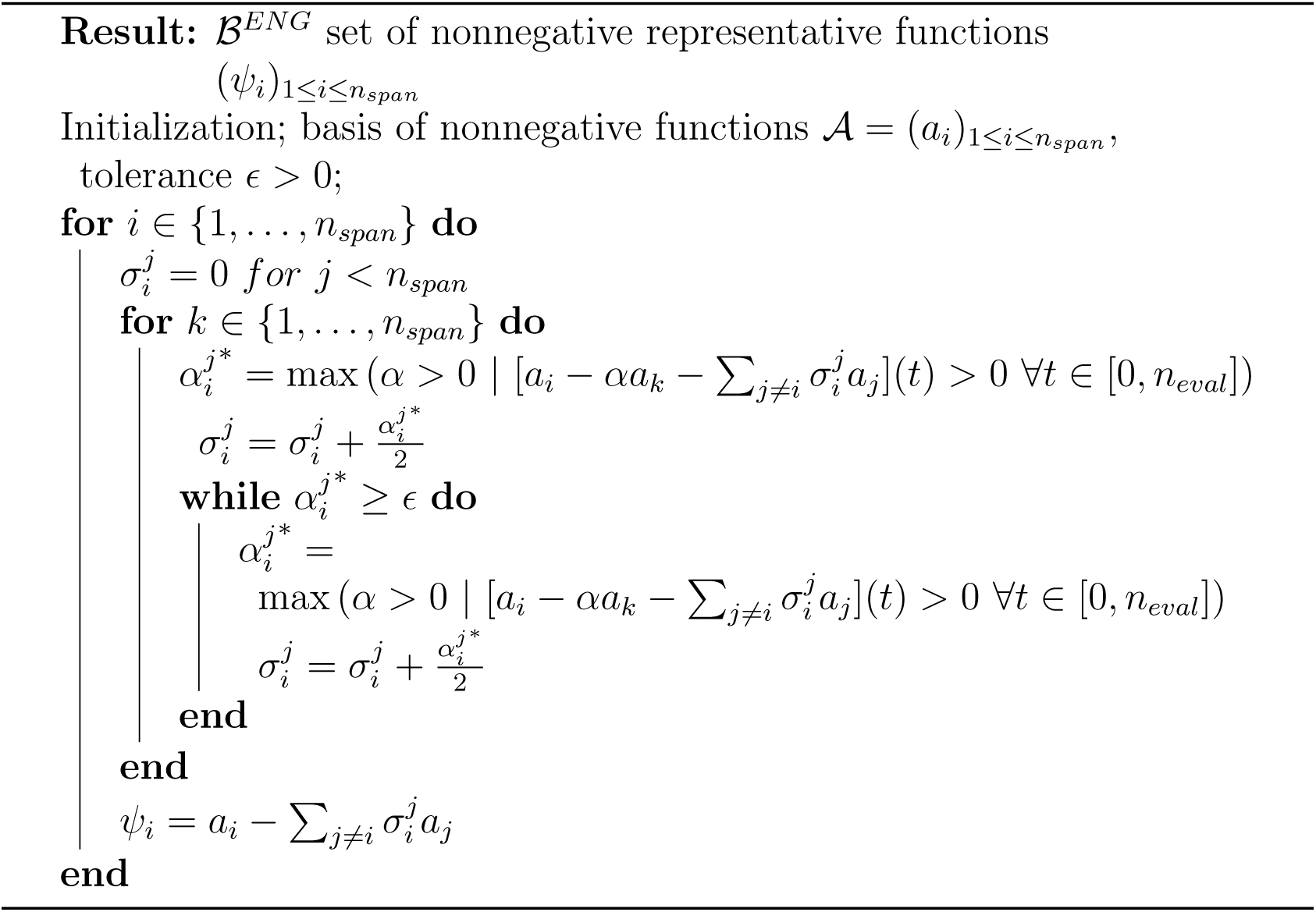

Note that, due to numerical integration errors, some of the curves in Figure 8 take a few values below zero. However in practice, applying Algorithm 1 to these functions, we obtained positive functions in ℬ^*ENG*^ as can be seen on Figure 11. The same happened for 𝒢^*ENG*^ (defined from ***G*** as ℬ^*ENG*^ from ***B***).

**Figure 11:**
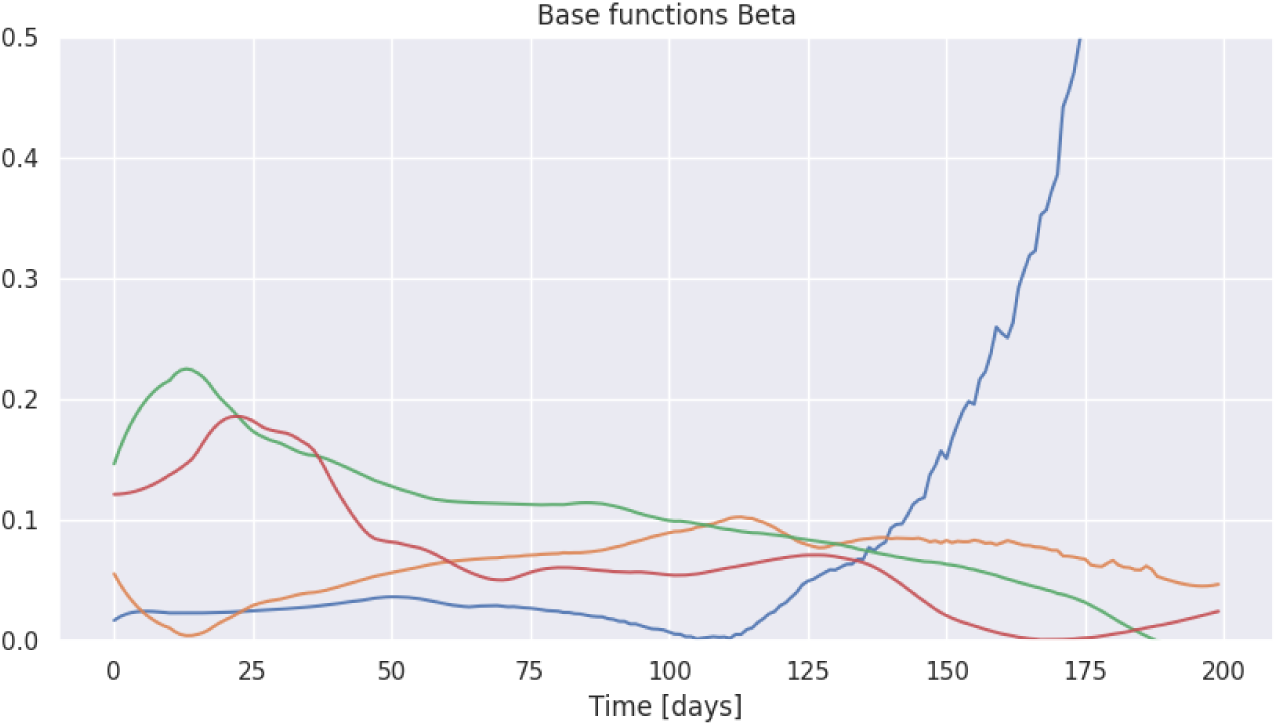
The set ℬ^*ENG*^ of functions obtained through the greedy algorithm combined with Algorithm 1 for *n*_*span*_ = 4.

**Figure 12:**
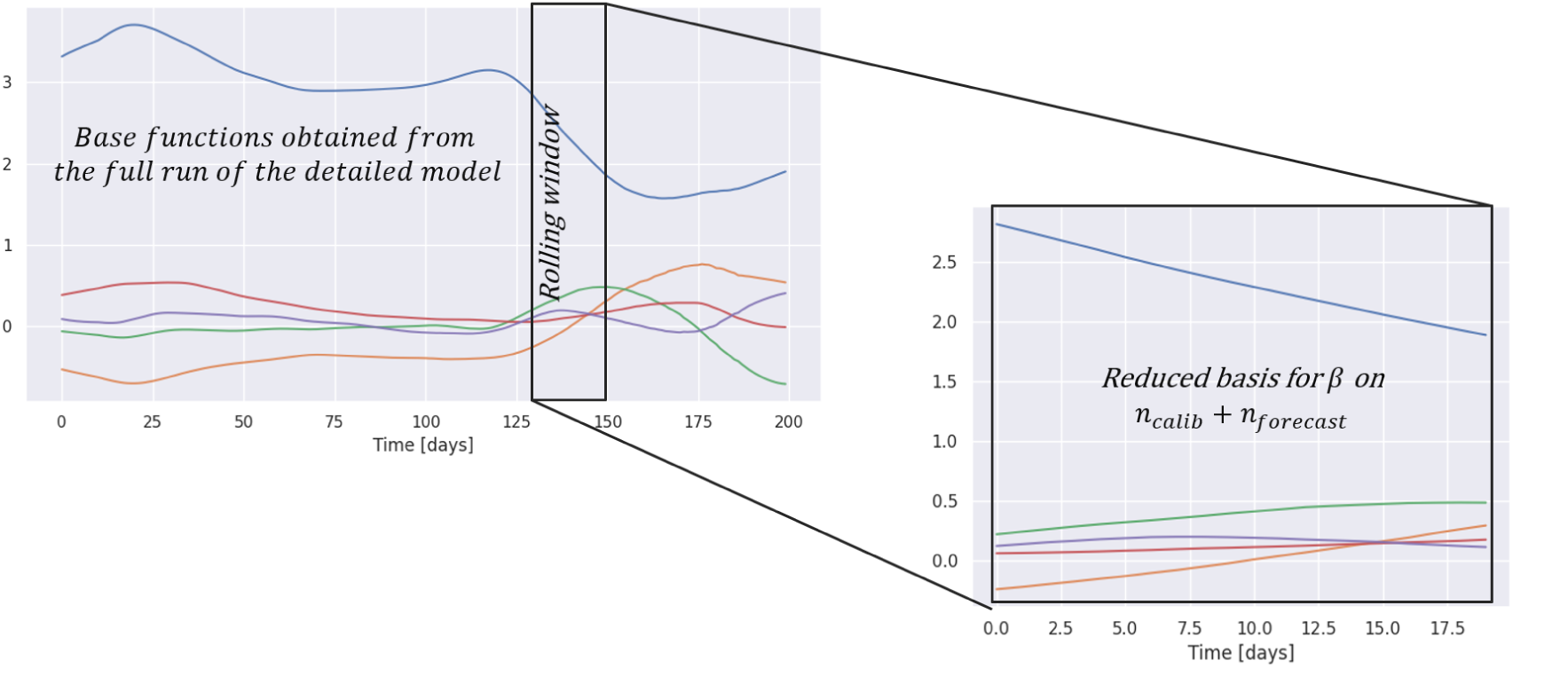
Extraction of a reduced basis from the functions obtained with the detailed model evaluations and the reduced order modeling algorithms presented in Sections 5.3.1 and 5.3.2.

### 5.4. Building a set of reduced bases for short-term forecasting by moving the initial time

In this section, we detail the way ℬ (obtained from SVD or ENG) was used to build a set of reduced bases for extrapolating 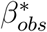.

#### 5.4.1. Extraction of a reduced basis

Given a set 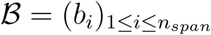 of functions defined on *n*_*eval*_ days, we extract a set of reduced bases (ℬ _*k*_)_1≤*k*≤*K*_ by applying a sliding window to these functions. More precisely, we first choose a window step *a* of 5 days. Then, for 1 ≤ *k* ≤ 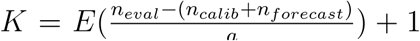, with *E*(*x*) the integer part of *x*, we denote ℬ_*k*_ the set of functions 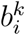 corresponding to the restriction of *b*_*i*_ to the window [(*k* − 1) × *a*, (*k* − 1) × *a* + *n*_*calib*_ + *n*_*forecast*_], for 1 ≤ *i* ≤ *n*_*span*_.

#### 5.4.2. Enrichment of the reduced bases

In Section 5.4.1 we built a set of reduced bases (B_*k*_)_1≤*k*≤*K*_ for extrapolating 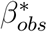 from the reduction of our detailed model evaluations. Additionally, we can add well-chosen functions to these reduced bases thanks to prior knowledge of the evolution of 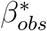. As can be seen on Figure 6, the variations of 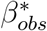 are mostly exponential. Starting at time *t*_0_ and given a *n*_*calib*_-day evolution of 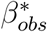, we can fit the following exponential function:

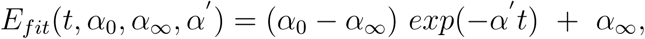

to 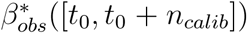 using least squares and add the function *E*_*fit*_([*t*_0_, *t*_0_ + *n*_*calib*_ + *n*_*forecast*_]) to the reduced basis. This is illustrated in Figure 13. In the following, this raw exponential extrapolation will be used as benchmark extrapolation of 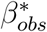 and 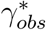 or in combination with the reduced bases obtained from ℬ^*SVD*^ and ℬ^*ENG*^.

**Figure 13:**
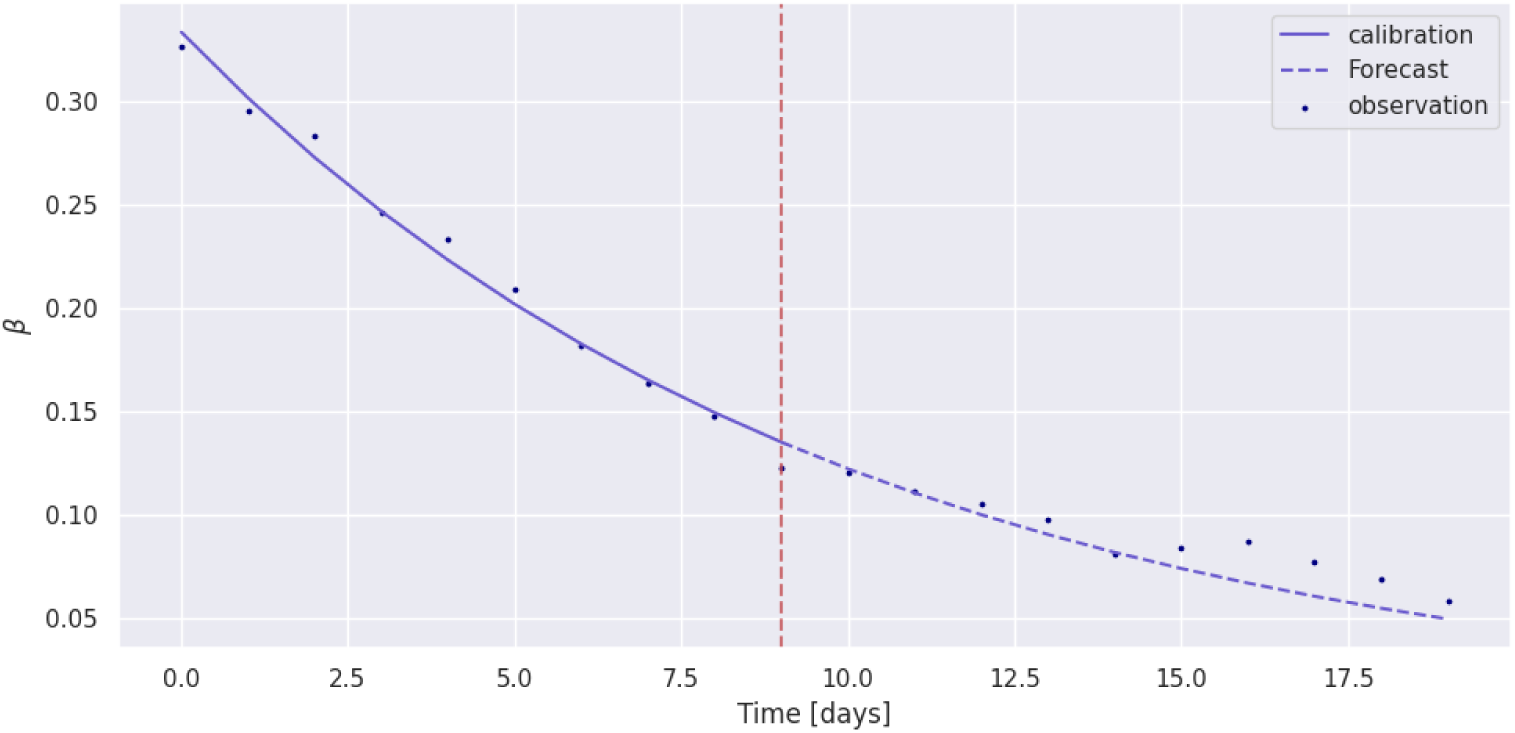
Exponential fit and forecast of the observed *β* in Isère department over a 20-day window starting on March 17^th^, 2020. Here *t*_0_ = 0, *n*_*calib*_ = 10, *n*_*forecast*_ = 10.

### 5.5. Short-term forecasts of the infection number

We now have *K* potential candidates for the reduced basis of 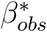 extrapolation. In this section, we present the way we selected the best reduced basis, and then how we used it to get a forecast on the infection number.

#### 5.5.1. Selection of the best forecast for beta

For computing a forecast on [*t*_0_ + *n*_*calib*_ + 1, *t*_0_ + *n*_*calib*_ + *n*_*forecast*_], we divided the calibration period in *T*_1_ = [*t*_0_, *t*_0_ + *n*_1_ − 1] and *T*_2_ = [*t*_0_ + *n*_1_, *t*_0_ + *n*_*calib*_]. Then for each reduced basis 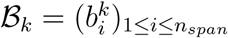, we defined the loss function:

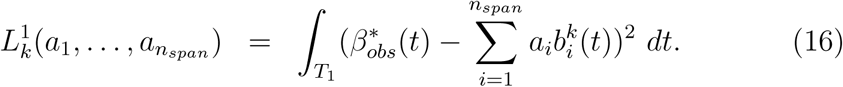

By optimizing it, we computed a function 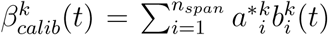 defined on [*t*_0_, *t*_0_ + *n*_*calib*_ + *n*_*forecast*_] fitting at best 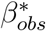 on *T*_1_. We then computed:

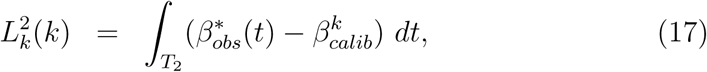

and selected the function 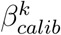 with the lowest evaluation of the loss 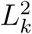 for extrapolating 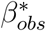 on [*t*_0_ + *n*_*calib*_ + 1, *t*_0_ + *n*_*calib*_ + *n*_*forecast*_].

#### 5.5.2. Short-term forecast of the infection number

By applying the methodology presented in Section 5.5.1 to the parameter *β*, we obtained a calibration-forecast function *β*_*calib*_ as presented on Figure 14. The methodology was applied in parallel to the parameter *γ*. From *β*_*calib*_ and *γ*_*calib*_, we could run the time-dependent SIR model defined by (1) with initial conditions (*I*_*obs*_(*t*_0_), *R*_*obs*_(*t*_0_)) and parameters *β* = *β*_*calib*_, *γ* = *γ*_*calib*_. The model output on the infection number is shown on Figure 15.

**Figure 14:**
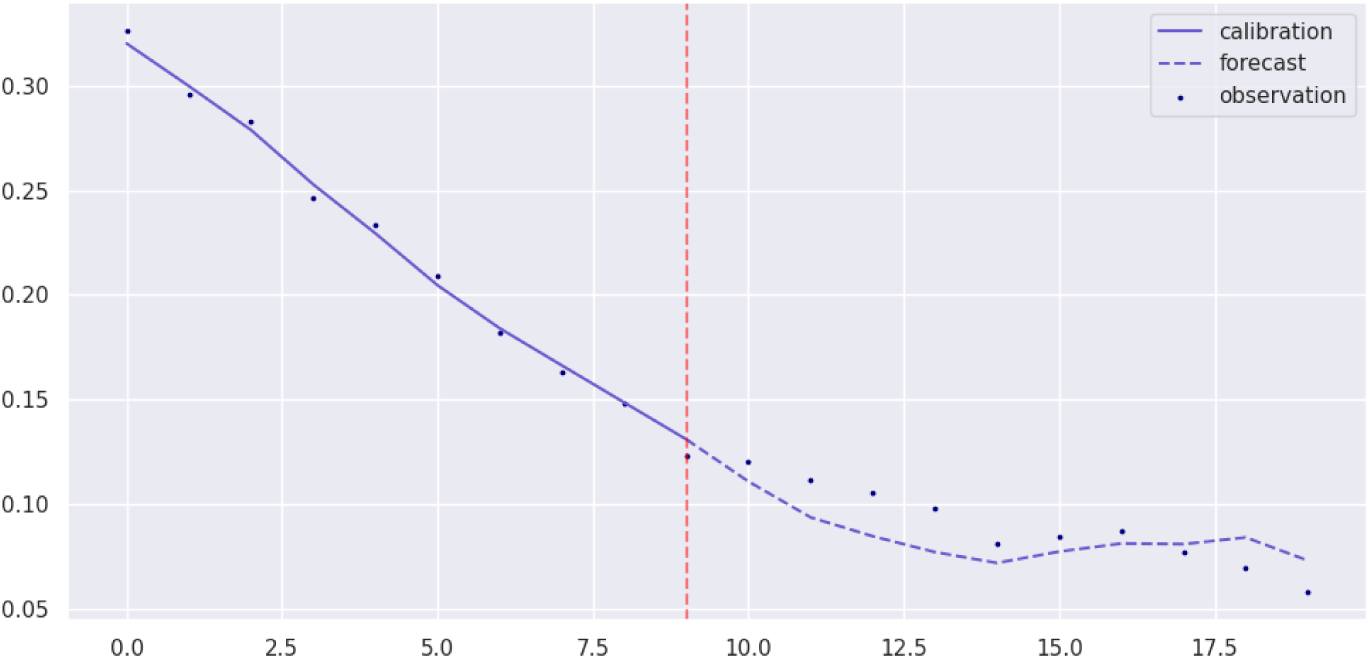
10-day fit followed by a 10-day forecast of 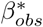 using the SVD basis presented in 5.3.1.

**Figure 15:**
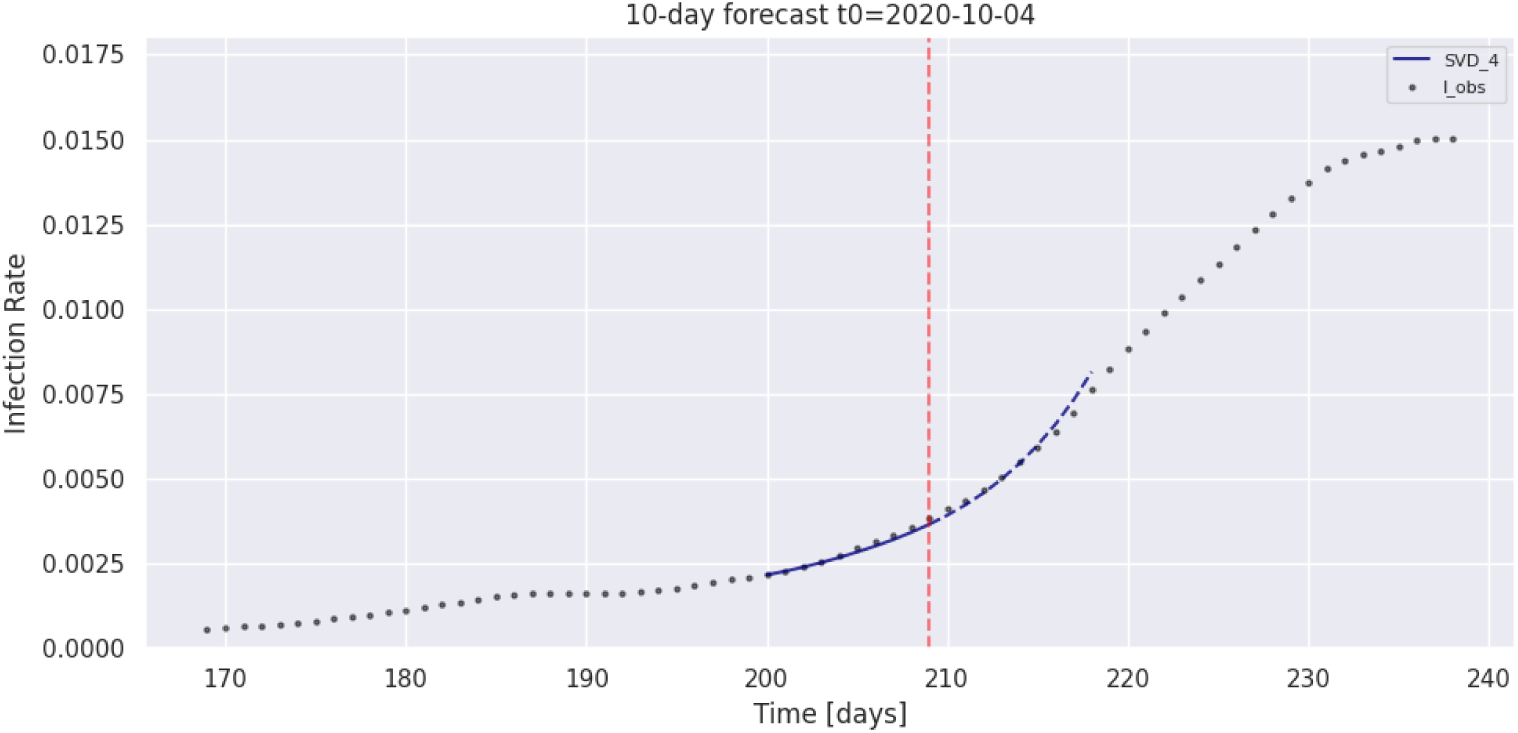
Forecast of the infection number in Isère department using the SVD reduced basis with *n*_*span*_ = 4.

### 5.6. Comparison of the SVD and greedy algorithms

To evaluate the error of our models, we generated 10-day forecasts every 5 days between March 2020 and May 2021. In this section and in the following, the forecast error for a calibration starting at *t*_0_ is:

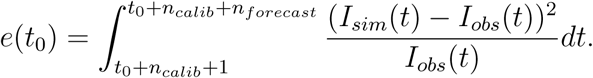

We compared the SVD and Greedy algorithms with and without the addition of an exponential extrapolation of 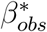, the number of basis elements was set to *n*_*span*_ = 5. The average error on the full timeframe are shown on Figure 16. The ENG and SVD algorithms performed very similarly, and the exponential enrichment of the reduced bases did not improve significantly the forecasting score. Therefore in the following we computed 10 day forecasts with the SVD algorithm, as it is much faster. Also, in the following sections, we did not use the exponential enrichment.

**Figure 16:**
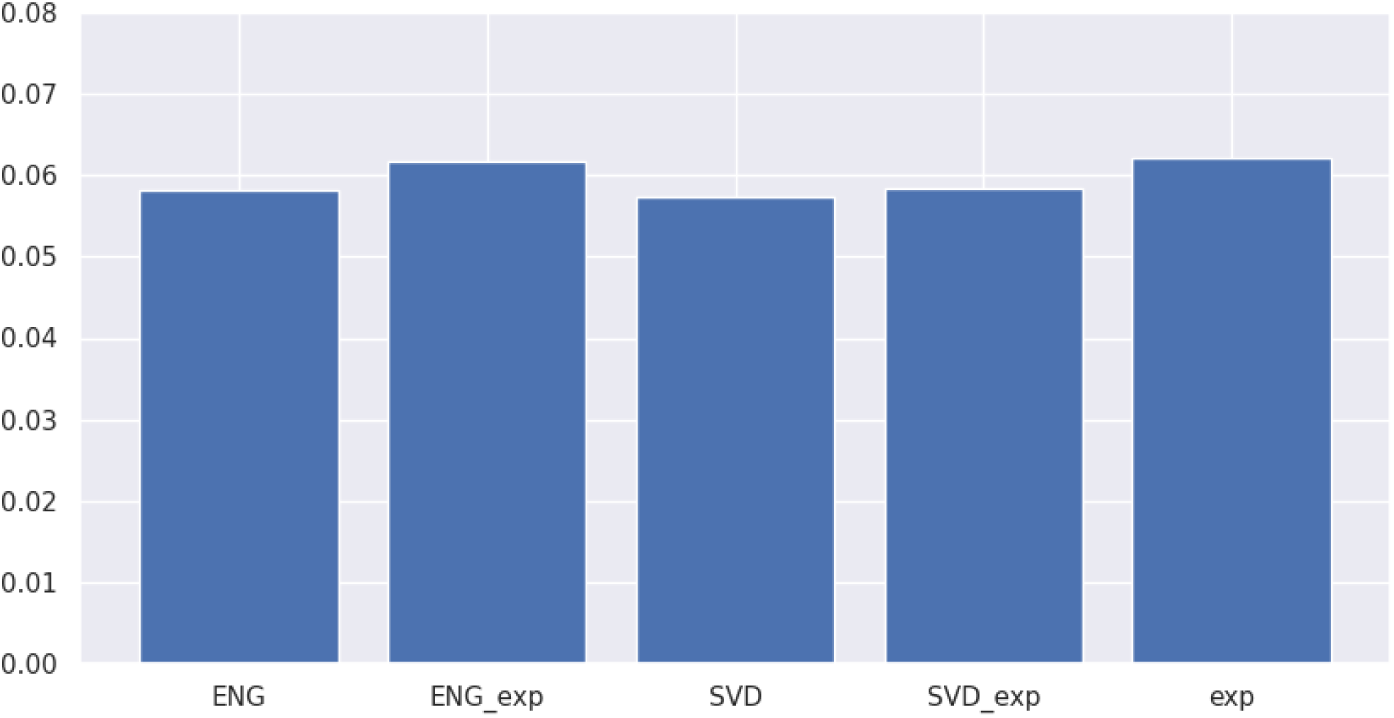
10-day forecast error for the greedy algorithm (*ENG*), the enriched greedy algorithm (*ENG*_*exp*_), the SVD algorithm (*SVD*), the enriched SVD algorithm (*SVD*_*exp*_) and the exponential benchmark algorithm (*exp*). We can see that the enrichment does not improve significantly the forecasting score, and that the SVD and ENG algorithms perform very similarly.

### 5.7. Building an aggregated model

In this section, we chose to focus on the SVD algorithm that we evaluated on 10-day forecasts. The choice of *n*_*span*_ is crucial because fewer basis functions may not be sufficient for capturing the complexity of the problem while a too large span of basis functions could lead to overfitting over the calibration and deteriorating the forecast accuracy of the reduced basis. We compared the models obtained by using different sizes of reduced basis, and built an aggregated model (*Agg*) that averages the forecasts obtained with the different basis size.

We computed the forecast errors on the infection number during a full year of 10-day forecasts with different sizes of the reduced basis, keeping the same number of basis functions for *β* and *γ*. The results are shown on Figure 17 for the SVD algorithm. We can see that our aggregated model performed better than any individual model, and that this model reached a 5.5% error on 10 day forecasts. The visualization of some individual model predictions can be seen on Figure 19. Moreover, the aggregated model seems more robust than the exponential extrapolation, preventing from very large errors as can be seen, e.g., on Figure 18.

**Figure 17:**
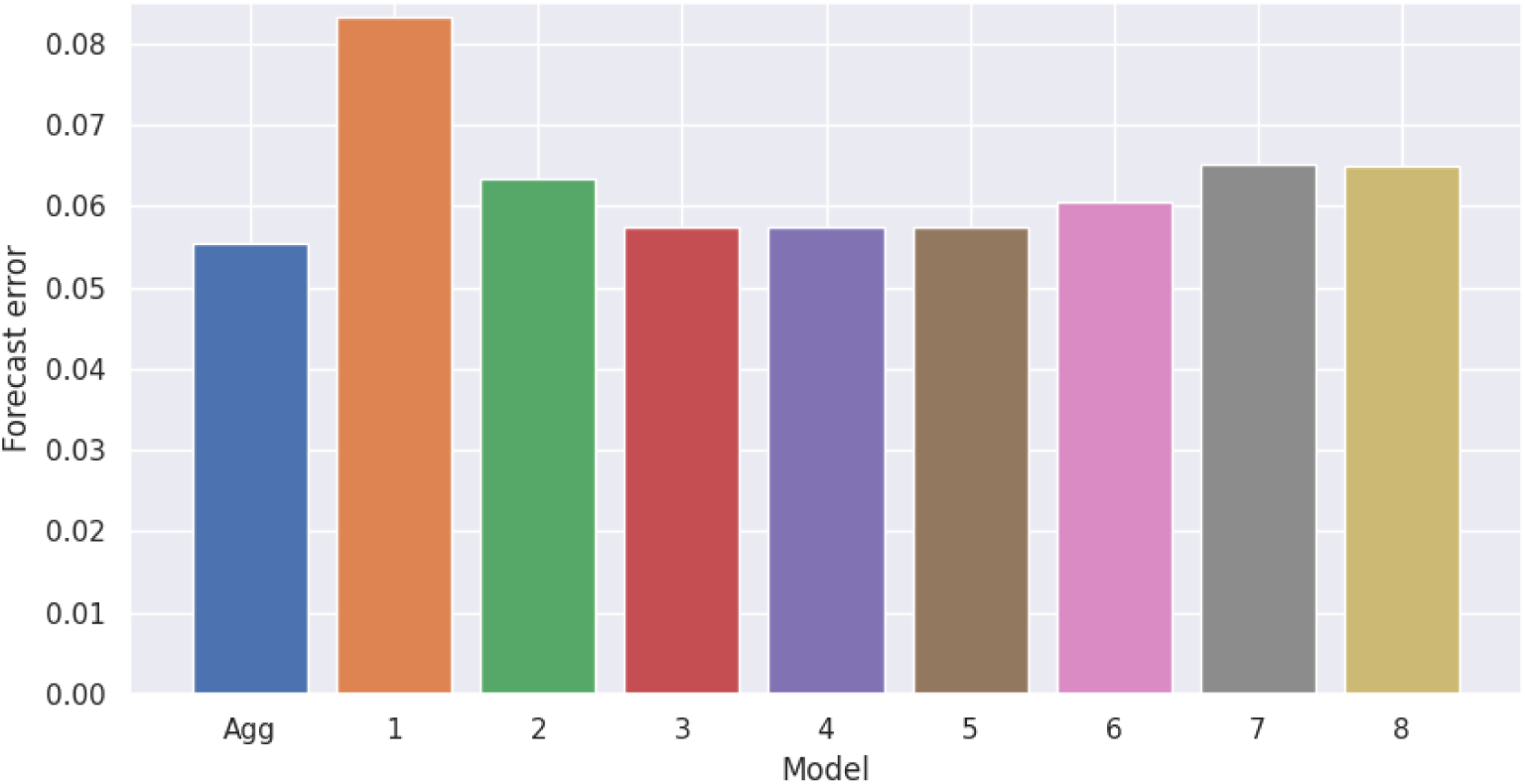
Forecast error of the aggregated model and of each individual model obtained with *n*_*span*_ ∈ [1, 8]. The aggregated model is the average of models 3, 4 and 5. We can see that the aggregated model performed slightly better than any of these individual models.

**Figure 18:**
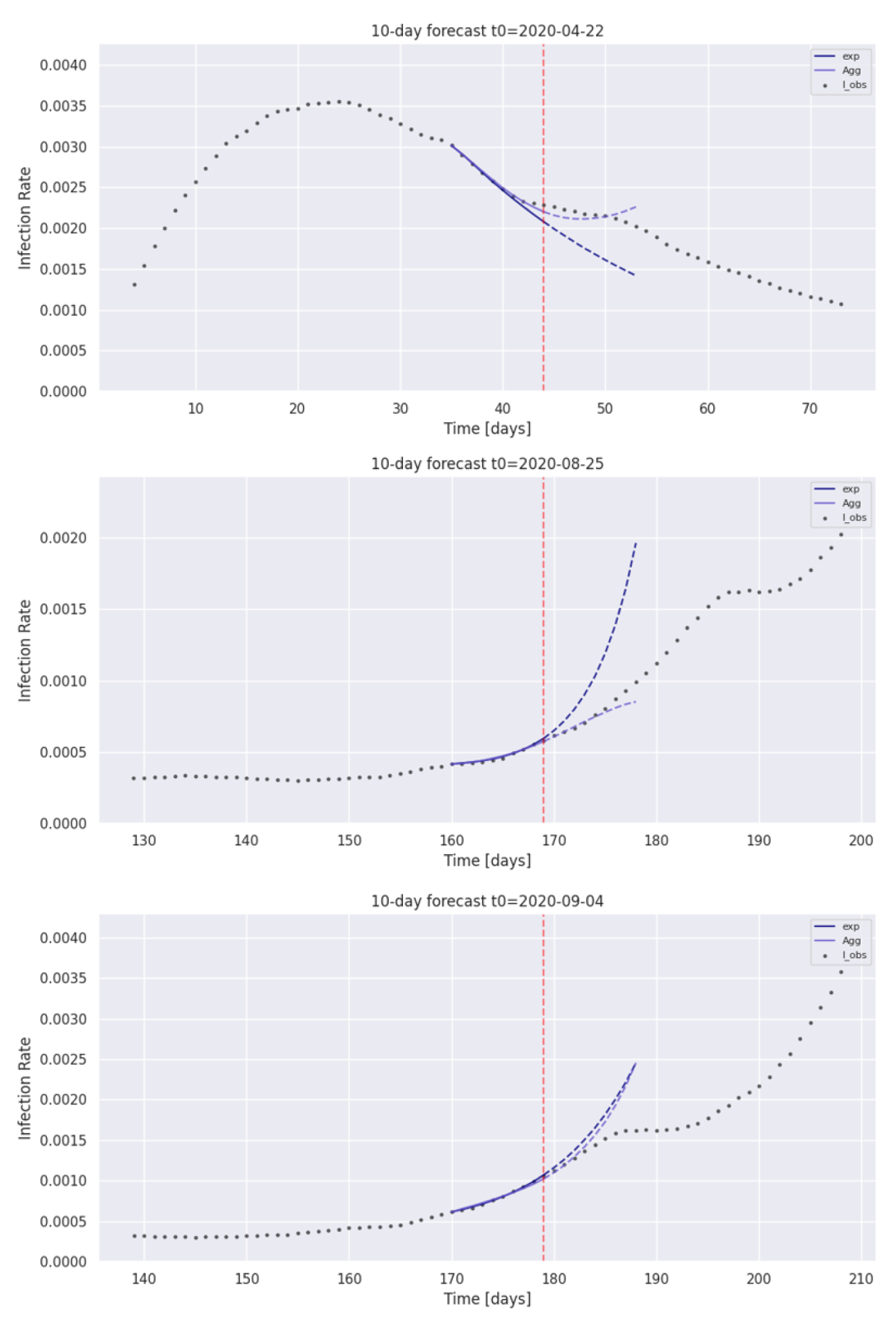
Comparison of forecasts obtained with the aggregated and exponential models at periods where the dynamics of Covid-19 changed quickly. We can see that the exponential extrapolation of 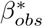 is more prone to exploding forecasts while the aggregated model has a more robust behaviour. Additionnaly, the third plot shows an example of a change in the dynamics that creates a large error as the forecasts can not anticipate a brutal change, e.g., in sanitary restrictions.

**Figure 19:**
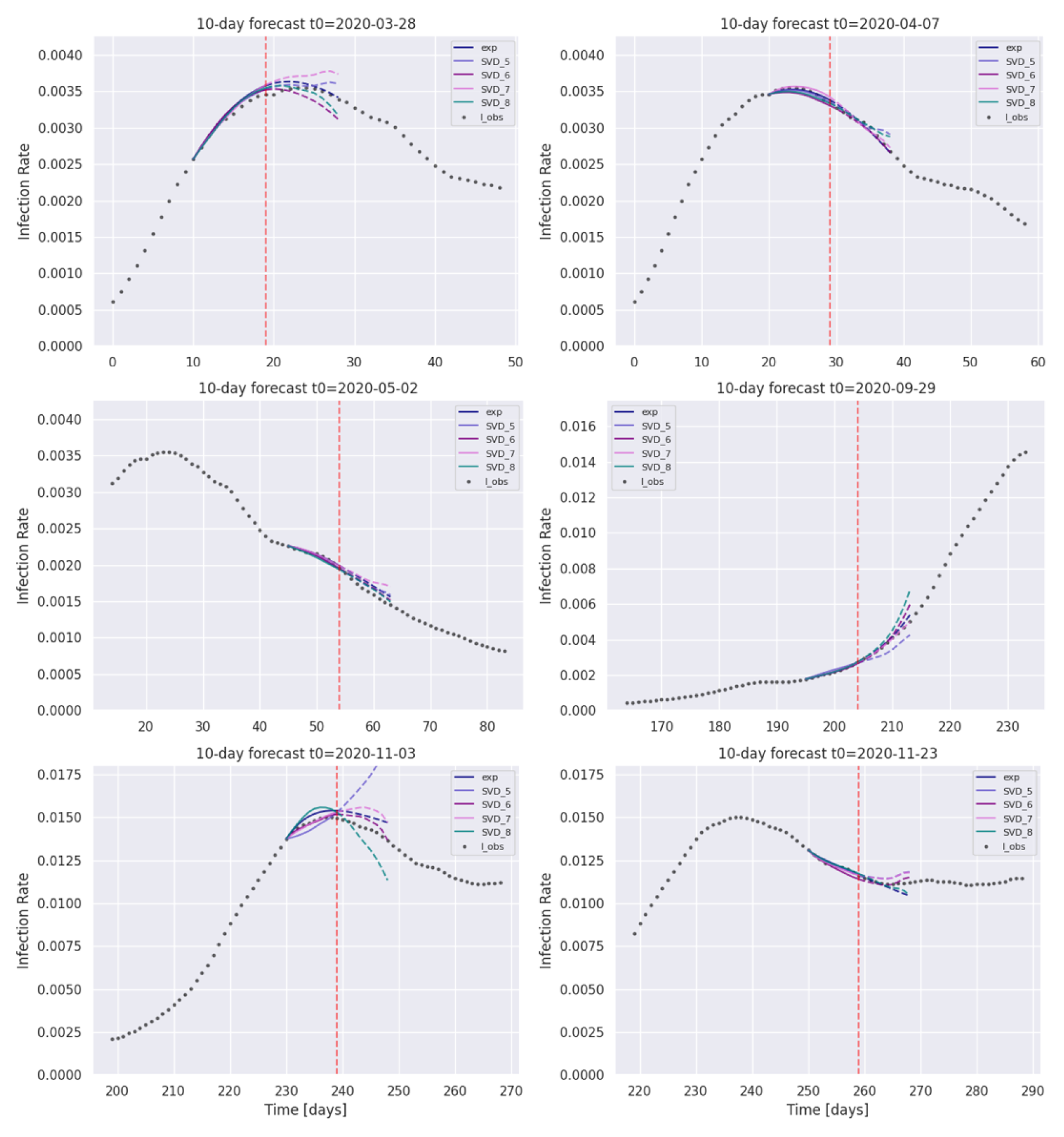
Visualization of the individual model forecasts in different phases of the pandemic. The bottom left plot illustrates the fact that the aggregated model performs better than any individual model.

### 5.8. Epidemiological forecast using the detailed model

To evaluate the efficiency of the reduced modeling approach for forecasting, we compared its cost with the one consisting in forecasting the infection number directly from the detailed model. All the computation times mentioned below were obtained on a regular laptop. In the reduced basis approach, the offline phase consists of *n*_*runs*_ = 500 evaluations of the detailed model over *n*_*days*_ = 200 days. The detailed model runs in *τ* = 0.15 *s.day*^−1^, which totals *C*_off_ = *n*_*runs*_ *n*_*days*_ *τ* = 15000 seconds. The online phase has a cost of *C*_*on*_ = 26 seconds, which is the time for projecting 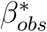 and 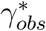 onto the reduced bases (ℬ _*k*_)_1≤*k*≤*K*_ and (𝒢 _*k*_)_1≤*k*≤*K*_, as described in 5.5.1. The cost of running the reduced model itself is negligible because it is run only once, with the optimal extrapolations of *β* and *γ*. Hence, the total cost of the reduced model approach for *κ* forecasts is *κ C*_*on*_ + *C*_*off*_.

In the direct approach, the detailed model is evaluated over *n*_*calib*_+*n*_*forecast*_ = 20 days and the optimization of the 9 parameters presented in Section 5.2 required *n*_*eval*_ = 600 evaluations, totalling a direct time of *C* = (*n*_*calib*_ + *n*_*forecast*_) × *n*_*eval*_ × *τ* = 1800 seconds per forecast.

For *κ* forecasts, the gain is 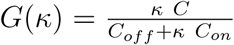 With *κ* ≈ 10 forecasts, the cost of the offline phase is compensated by the gain in the online phase.

### 5.9. Discussion

This study has shown that extrapolating 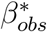 and 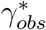 using the exponential function as presented in 5.4.2 is a reasonable solution for forecasting the infection number at a 10-day timescale. However our approach led to a slightly better forecasting score with a model that is more robust, hence more reliable. We would like to point out that the forecasting score has an underlying error due to the changes in dynamics of the epidemic with sanitary measures that are taken. In fact, the model has large overestimations of the infection number every time a new measure is taken, as can be seen on the bottom plot of Figure 18. As a consequence, the forecasting error could not drop to a very low value without inputting the sanitary measures into the model. The model would perform better with no changes in the dynamics, however this model is still very useful because it gives the evolution of the pandemic if no measures were taken, which can help to measure the efficiency of the restriction rules.

## 6. Conclusion

In this paper, we presented a spatialized extension of a detailed mutli-compartmental epidemiological model that allowed us to reproduce the evolution of hospital needs at the scale of the department of Isère during the Covid-19 outbreak, while giving a detailed information about the geographical heterogeneity of the sanitary conditions. The partial differential equations that define the model were solved with a very efficient meshless scheme on a very irregular domain. From this model we built a reduced basis for extrapolating the transmission rate *β* and recovery rate *γ* obtained from hospital data and we used them in a surrogate model to output forecasts of the global infection number in the department of Isère. A simple exponential extrapolation proved to be efficient for the extrapolation of the transmission and recovery rates but the aggregation of surrogate models using different reduced bases gave a more robust forecast. The work presented in the present paper could be used to evaluate the efficiency of restriction rules that are taken by comparing the forecasts as a reference for the evolution of the pandemic with the actual evolution of the infection number when restriction rules are taken. Also, testing policies could be evaluated by controlling parameters *λ*_1_ and *λ*_2_ and by evaluating the dynamics of the pandemic while more people are being isolated after being tested. Note that the spatialized detailed model we introduced could be completed to account, e.g., for vaccination or emergence of a variant. We could add compartments for the vaccinated population by calibrating their own probability of transmission and severity of symptoms. In particular, this could allow to compare different vaccination policies. The spatialized model could take into account age categories using contact, testing and vaccination rates as well as symptom severity in each age category. This could be solved with the same approach, but would benefit from more precise hospital data involving the age of patients for calibration.

## Data Availability

All data produced in the present study are available upon reasonable request to the authors

